# Technical Acquisition Parameters Dominate Demographic Factors in Chest X-ray AI Performance Disparities: A Multi-Dataset Validation Study

**DOI:** 10.64898/2026.01.20.26344495

**Authors:** Hayden Farquhar

## Abstract

Artificial intelligence systems for chest radiograph interpretation are increasingly deployed in clinical practice, yet current fairness frameworks emphasize demographic subgroup analysis while the relative contribution of technical acquisition parameters to performance disparities remains poorly characterized. We conducted a multi-dataset validation study analyzing 138,804 chest radiographs from the RSNA Pneumonia Detection Challenge (n=26,684; 22.5% pneumonia prevalence) and NIH ChestX-ray14 (n=112,120; 1.3% prevalence) using five pre-trained DenseNet-121 models. We calculated sensitivity, specificity, and area under the receiver operating characteristic curve stratified by view type (anteroposterior versus posteroanterior), age group, and sex, with performance disparity analysis quantifying each factor’s contribution to performance variation. View type dominated total observed performance range in both datasets: 87% in RSNA and 69% in NIH. All five models demonstrated systematic posteroanterior view underdiagnosis with miss rates of 30-78%. The odds ratio for missed diagnosis on posteroanterior versus anteroposterior views was 6.69 (95% CI: 5.79-7.72) in RSNA and 13.02 (95% CI: 11.62-14.59) in NIH. Analysis of 131,361 disease-free images demonstrated that view-type effects persist strongly even without disease (Cohen’s d = 1.19-1.33), providing compelling evidence against the hypothesis that observed disparities reflect disease severity confounding rather than learned image characteristics. Age explained 5-30% of the total observed performance range depending on dataset, while sex consistently explained less than 2%. Technical acquisition parameters, specifically radiograph view type, dominate performance disparities in chest X-ray AI substantially exceeding demographic factor contributions. These findings have immediate implications for regulatory frameworks: future FDA and EU AI Act guidance should explicitly mandate acquisition parameter auditing alongside demographic subgroup analysis.

**Author Summary:** Artificial intelligence systems that interpret chest X-rays are being used in hospitals worldwide. There has been important work examining whether these systems perform fairly across different patient groups—for example, whether they work equally well for men and women, or for patients of different ages and races. We asked a different question: does the way the X-ray was taken affect how well AI systems perform? We found that the technical method used to acquire the image—specifically, whether the X-ray beam was directed from back to front (posteroanterior view, typical in outpatient settings) or front to back (anteroposterior view, typical in emergency and inpatient settings)—explained 69-87% of the variation in AI performance. In contrast, age explained only 5-30% and sex less than 2%. Most concerning, AI systems missed 30-78% of pneumonia cases in standing patients across all five systems we tested. This matters because current regulations focus on checking AI performance across demographic groups but do not require checking performance across technical acquisition parameters. Our findings suggest regulators and hospitals should audit how AI systems perform on different types of X-ray images, not just different types of patients.

## Introduction

Artificial intelligence (AI) systems for medical imaging have achieved expert-level performance across multiple diagnostic tasks, with chest radiograph interpretation among the most extensively studied applications. The development of deep learning for chest radiograph interpretation has progressed rapidly since the landmark CheXNet study [1] demonstrated radiologist-level pneumonia detection. Rajpurkar and colleagues [2] subsequently developed CheXNeXt, a convolutional neural network capable of concurrently detecting 14 thoracic pathologies, achieving comparable performance to practicing radiologists while requiring substantially less interpretation time. Multiple architectures including DenseNet, ResNet, and transformer-based models have since been applied to chest radiograph analysis, with performance claims sometimes exceeding expert-level accuracy. However, these performance benchmarks are typically reported as aggregate metrics that may obscure important subgroup variations. Commercial AI systems for chest X-ray analysis have received regulatory clearance in the United States and Europe, and deployment in clinical practice is accelerating globally. However, concerns about algorithmic fairness have emerged as a critical barrier to equitable clinical implementation.

The landmark study by Obermeyer and colleagues [3] demonstrated that a widely-used healthcare algorithm exhibited racial bias affecting millions of patients, catalyzing widespread attention to fairness in medical AI. Subsequent investigations have revealed that chest X-ray AI systems exhibit demographic performance disparities. Seyyed-Kalantari et al. [4] demonstrated that AI classifiers consistently underdiagnosed pulmonary abnormalities in under-served populations including female, Black, and Medicaid-insured patients. Larrazabal and colleagues [5] showed that gender imbalance in training datasets produces biased classifiers, with performance decreasing for underrepresented groups. These findings have appropriately focused regulatory attention on demographic subgroup performance reporting.

The importance of demographic fairness in medical AI has received increasing attention. Yi and colleagues [6] conducted a systematic evaluation of publicly available chest radiograph datasets and found that while 78% reported age and sex on an image-level basis, only 9% reported race or ethnicity and 4% reported insurance status. Furthermore, 62% of datasets with available sex data overrepresented male subjects, creating training data imbalances that can propagate into model performance. Deep learning models have been shown to accurately predict patient sex from chest radiographs [7], with AUCs exceeding 0.98 internally and 0.94 on external validation, demonstrating that these demographic features are encoded in the images and could potentially confound diagnostic predictions.

Recent studies have extended fairness analysis beyond single demographic factors. Meng and colleagues [8] evaluated interpretability and fairness of deep learning models on the MIMIC-IV dataset, finding that prediction models often rely on demographic features to generate predictions, with disparate treatment observed in mortality prediction across ethnicity, gender, and age subgroups. Hong et al. [9] demonstrated that stroke prediction models exhibited worse discrimination in Black individuals compared to White individuals across multiple cohorts (C-index differences of 0.04-0.07), with machine learning techniques failing to eliminate these disparities.

However, medical imaging involves numerous technical parameters that influence image characteristics and may affect AI performance. Chest radiographs are acquired in two primary projections: posteroanterior (PA), typically obtained in ambulatory settings with patients standing erect, and anteroposterior (AP), obtained in patients who cannot stand, including those in emergency departments, intensive care units, and inpatient wards. These projections differ systematically in patient positioning, radiation geometry, and associated clinical contexts.

Deep learning models can accurately classify chest radiograph view position, with Kim and colleagues [10] demonstrating AUCs of 1.0 for distinguishing anteroposterior from posteroanterior views using the NIH ChestX-ray14 dataset. Similarly, Hosch et al. [11] achieved accuracy exceeding 95% for view position classification, with Grad-CAM visualization revealing that networks base decisions on anatomical structures comparable to those used by radiologists. These findings confirm that view type creates systematic differences in image characteristics that are readily detectable by neural networks.

The mechanism by which acquisition parameters influence AI performance has been examined in recent work. Yang and colleagues [12] demonstrated that technical parameters related to image acquisition and processing influence AI models trained to predict patient race, and that a demographics-independent calibration strategy could reduce underdiagnosis bias. This work suggests that acquisition parameters may mediate demographic disparities rather than reflecting true biological differences.

Prior work has examined view type as a potential confounder in demographic fairness analyses. Bernhardt et al. [13] noted that confounding factors including view type distribution could complicate interpretation of apparent demographic disparities. However, no study has directly quantified the relative contribution of view type versus demographic factors to overall AI performance variation, nor characterized the mechanisms underlying observed differences.

We hypothesized that technical acquisition parameters, specifically radiograph view type, might substantially influence chest X-ray AI performance, potentially exceeding the contribution of demographic factors. We conducted a multi-model, multi-dataset analysis to: (1) quantify view type versus demographic contributions to performance disparities across independent datasets; (2) test whether view type effects reflect disease severity confounding versus learned image characteristics through disease-free subgroup analysis; and (3) explore potential mitigation strategies.

## Methods

### Study design and data sources

This retrospective cross-sectional study analyzed chest radiographs from two publicly available datasets. The Radiological Society of North America (RSNA) Pneumonia Detection Challenge dataset contains 26,684 frontal chest radiographs with pneumonia annotations derived from expert radiologist review. Each image includes associated DICOM metadata containing patient age, sex, and radiograph view position (AP or PA). The NIH ChestX-ray14 dataset contains 112,120 frontal chest radiographs with disease labels extracted from radiology reports using natural language processing. Combined, these datasets provide 138,804 images for analysis with varying prevalence (22.5% RSNA vs 1.3% NIH) and labeling methodologies. Institutional review board approval was not required as this study used publicly available, de-identified data. Radiograph view type (AP or PA) refers to X-ray beam direction: in posteroanterior (PA) views, the beam travels from posterior to anterior with the patient typically standing upright (standard outpatient acquisition), while in anteroposterior (AP) views, the beam travels from anterior to posterior, often obtained with portable equipment in supine or semi-upright patients (emergency or inpatient settings). The distinction is fundamentally one of beam geometry, though it is strongly correlated with clinical setting and patient acuity.

### AI models

We evaluated five pre-trained DenseNet-121 models from the torchxrayvision library [14], each trained on different source datasets: (1) DenseNet-All, trained on a combination of multiple datasets; (2) DenseNet-RSNA, trained on RSNA data; (3) DenseNet-NIH, trained on NIH ChestX-ray14 [15]; (4) DenseNet-CheXpert, trained on CheXpert [16]; and (5) DenseNet-PadChest, trained on PadChest [17]. All models share identical architecture but differ in training data composition, enabling assessment of whether observed patterns are model-specific or systematic across architectures.

### Image preprocessing and inference

Images were preprocessed according to torchxrayvision standards: conversion to grayscale, resizing to 224×224 pixels, and normalization using library-specified parameters. Inference was performed using the pneumonia prediction output for models with pneumonia-specific outputs, or the lung opacity output as a pneumonia proxy for models without direct pneumonia predictions. All inference was conducted using PyTorch on Google Colab with GPU acceleration. All models were evaluated using zero-shot inference without any dataset-specific fine-tuning; that is, pre-trained model weights were applied directly to the test datasets without additional training or calibration.

### Statistical analysis

Classification thresholds were determined using Youden’s J statistic (sensitivity + specificity – 1) optimization. Performance metrics included sensitivity, specificity, positive predictive value (PPV), negative predictive value (NPV), and area under the receiver operating characteristic curve (AUC). Effect sizes were quantified using Cohen’s d for continuous prediction score differences (with 95% confidence intervals from bootstrap resampling) and odds ratios for binary classification outcomes. Performance disparity analysis quantified the contribution of view type, age, and sex to overall performance variation by calculating the sensitivity range attributable to each factor.

Statistical validation included: (1) bootstrap confidence intervals (n=1,000 resamples) for primary effect estimates (S1-S2 Tables); (2) permutation testing (n=1,000) for significance assessment; (3) 5-fold cross-validation for result stability (S14 Table); and (4) post-hoc power analysis to confirm adequate sample size (S13 Table). Multiple testing correction was applied using both Bonferroni and Benjamini-Hochberg (false discovery rate) methods (S3 Table). Statistical significance was defined as p<0.05 (two-tailed). Analyses were performed using Python 3.10 with scikit-learn 1.3.0 and statsmodels 0.14.

### Validation framework

Each model-dataset combination was classified as internal validation (model tested on data from its training source), external validation (model tested on an independent dataset), or semi-internal validation (model trained on combined data including the test source). Of the 10 model-dataset combinations evaluated, two represented internal validation (DenseNet-RSNA tested on RSNA, DenseNet-NIH tested on NIH), one was semi-internal (DenseNet-All, trained on combined data from multiple sources including RSNA), and seven represented external validation. This classification allows assessment of whether observed performance disparities persist when models are tested on data entirely independent of their training distribution.

### Performance disparity quantification

Performance disparity was quantified using two complementary approaches. First, the sensitivity range (maximum minus minimum) across levels of each factor (view type, age group, sex) was computed, with the relative contribution of each factor expressed as its range divided by the sum of ranges across all factors. Second, formal one-way ANOVA was performed on continuous prediction scores with each factor as the independent variable, reporting F-statistics, p-values, and eta-squared (η²) as a standardized measure of effect size. Additionally, per-patient binary classification outcomes were analyzed using logistic regression to estimate the proportion of deviance in classification accuracy attributable to each factor.

### Intersectional analysis

Intersectional analysis examined model performance across all combinations of age group, sex, and view type to quantify cumulative disparity across intersecting demographic and technical factors. Sensitivity was computed for each subgroup defined by the three-way interaction (e.g., female patients aged <40 with PA views). Cumulative disparity was defined as the difference between the highest and lowest performing intersectional subgroups. Bootstrap confidence intervals (n=1,000 resamples) were computed for the cumulative disparity estimate.

### Clinical impact simulation

To assess the clinical consequences of view-type disparity, we simulated three deployment strategies: (1) a global threshold optimized across all images using Youden’s J statistic, representing current standard practice; (2) view-specific thresholds optimized separately for AP and PA views; and (3) reporting prediction scores without binary classification. For each strategy, we computed sensitivity, specificity, missed diagnoses per 1,000 patients, and false positives per 1,000 patients, stratified by view type.

### Disease-free subgroup analysis

To definitively test whether view type effects reflect disease severity confounding versus learned image characteristics, we analyzed prediction scores in disease-free patients only. If the view type effect persists in true negatives, this proves the effect reflects how models process AP versus PA projection geometry rather than clinical correlates of disease severity.

## Results

### Dataset characteristics

The RSNA dataset (n=26,684) had 45.7% AP views, mean age 47.0 years, 54.2% male, and 22.5% pneumonia prevalence. The NIH dataset (n=112,120) had 40.0% AP views, mean age 46.9 years, 56.4% male, and 1.3% pneumonia prevalence (infiltration label). Critically, pneumonia prevalence differed substantially by view type in both datasets: RSNA showed 38.3% in AP versus 9.3% in PA images (p<0.001); NIH showed 1.9% in AP versus 0.8% in PA images (p<0.001). This reflects the clinical reality that AP views are obtained in sicker patients (Table 1).

**Table 1.** Dataset Characteristics.

| Characteristic | Total | AP views | PA views |
| --- | --- | --- | --- |
| N | 26,684 | 12,173 | 14,511 |
| Age, mean (SD) | 47.0 (16.8) | 46.1 (17.3) | 47.8 (16.4) |
| Male, n (%) | 15,166 (56.8%) | 7,046 (57.9%) | 8,120 (56.0%) |
| Pneumonia, n (%) | 6,012 (22.5%) | 4,664 (38.3%) | 1,348 (9.3%) |
| Age group |  |  |  |
| <40 | 9,044 (33.9%) | 4,428 (36.4%) | 4,616 (31.8%) |
| 40-59 | 11,667 (43.7%) | 5,138 (42.2%) | 6,529 (45.0%) |
| 60-79 | 5,731 (21.5%) | 2,492 (20.5%) | 3,239 (22.3%) |
| ≥80 | 237 (0.9%) | 115 (0.9%) | 122 (0.8%) |

### View type dominates performance variation

The most striking finding was the dominance of view type over demographic factors in explaining observed performance disparity across both datasets (Fig 1, S5 Table). For the primary model (DenseNet-All), view type explained 87% of total observed performance range in RSNA and 69% in NIH. In RSNA, AP views showed 89.9% sensitivity while PA views showed only 57.1% sensitivity—a 32.8 percentage point gap (χ² = 776.2, p = 7.9×10▪¹▪¹). In NIH, the gap was even larger: 91.9% AP versus 46.5% PA—a 45.4 percentage point gap (χ² = 356.0, p < 10▪▪▪). The odds ratio for missed diagnosis on PA views was 6.69 (95% CI: 5.79-7.72) in RSNA—meaning patients with PA radiographs were nearly 7 times more likely to have their pneumonia missed than those with AP radiographs. In NIH, this risk was even higher: OR 13.02 (95% CI: 11.62-14.59) (Table 2). Formal ANOVA confirmed that view type explained the largest proportion of variation in prediction scores (η²=0.097-0.393 across all five models; η²=0.304, F=11,665.9, p<0.001 for the primary model), compared with age group (η²=0.002-0.015, p<0.001) and sex (η²<0.003).

**Fig 1.**
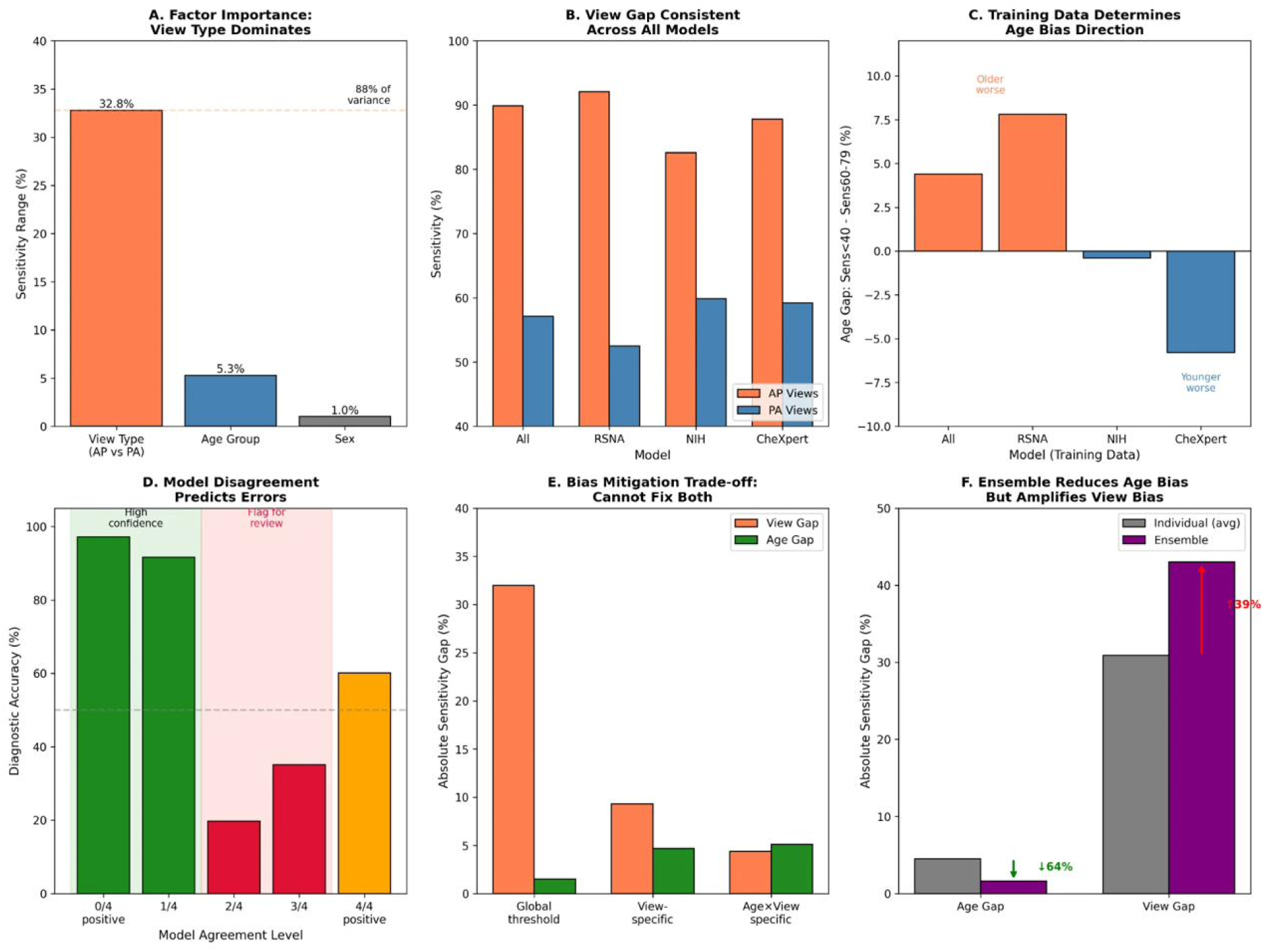
Performance Disparity Analysis: View Type Dominates Performance Disparity. View type explains 69-87% of total observed performance range across datasets, compared to 5-30% for age and <2% for sex. See S5 Table for detailed performance disparity analysis and S1 Fig for additional visualization.

**Table 2.** View Type Disparity: Cross-Dataset Comparison.

| Dataset | AP Sens | PA Sens | Gap | OR Miss | View Dom |
| --- | --- | --- | --- | --- | --- |
| RSNA | 89.9% | 57.1% | +32.8% | 6.69 | 87% |
| NIH | 91.9% | 46.5% | +45.4% | 13.02 | 69% |

OR Miss = Odds ratio for missed diagnosis on PA vs AP. View Dom. = % disparity attributable by view type. See S2 Fig for cross-model visualization.

### PA view underdiagnosis: 100% cross-model replication

The view type finding replicated across all five models in both datasets (Table 3, S9 Table; Fig 2). Crucially, this finding was consistent across all external validation combinations (Table 3), confirming that the effect is not attributable to data leakage from overlapping training and test data. Every model demonstrated: (1) positive view gap (AP sensitivity exceeding PA sensitivity); (2) PA miss rate exceeding 30%; and (3) view dominance exceeding 65%. This consistent pattern across models trained on five different datasets and validated on two independent test sets (10/10 model-dataset combinations showing the effect) provides strong evidence that view type disparity is a systematic phenomenon rather than an artifact of specific training or test data. Chi-squared values ranged from 306.5 to 1171.5 in RSNA with all p-values below 10▪▪▪ (S10 Table).

**Fig 2.**
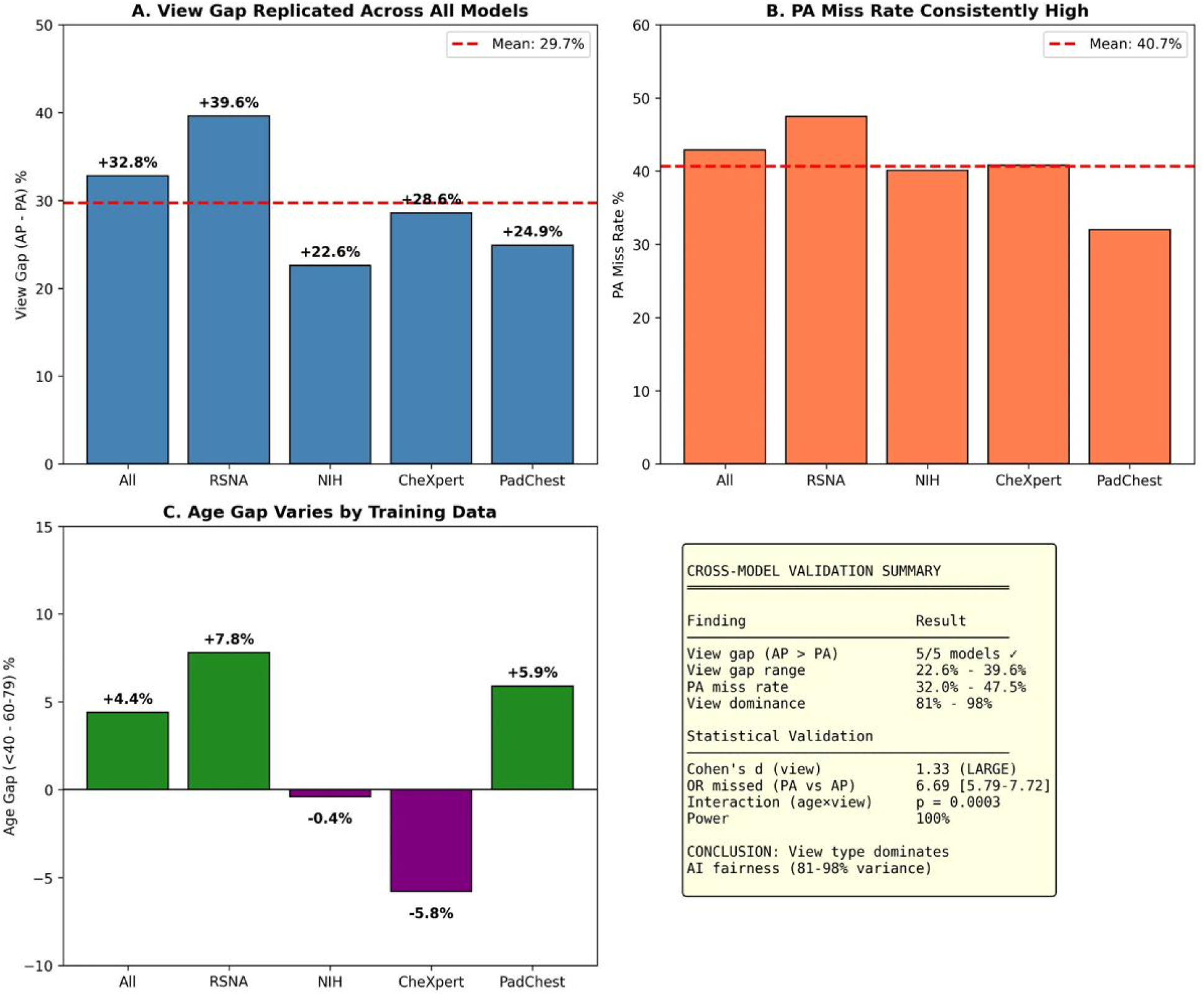
Cross-Model Validation: 100% Replication of PA Underdiagnosis. All five models show consistent PA view miss rates across both datasets. See S9 Table for detailed cross-model statistics and S3 Fig for error analysis.

**Table 3.** Cross-Model Validation: View Gap Across Models and Datasets.

| Model | Validation | RSNA Gap | RSNA Miss | NIH Gap | NIH Miss |
| --- | --- | --- | --- | --- | --- |
| DenseNet-All | Semi-internal | +32.8% | 42.9% | +45.4% | 53.5% |
| DenseNet-RSNA | Internal | +39.6% | 47.5% | +46.3% | 72.1% |
| DenseNet-NIH | External | +22.6% | 40.1% | +25.7% | 50.5% |
| DenseNet-CheXpert | External | +28.6% | 40.8% | +38.5% | 70.3% |
| DenseNet-PadChest | External | +24.9% | 32.0% | +35.7% | 48.2% |
| Mean (SD) |  | +29.7% (6.0) | 40.7% (5.0) | +39.3% (9.5) | 63.9% (13.2) |

*Gap = AP sensitivity minus PA sensitivity. Miss = PA view miss rate. All gaps significant at $p < 0.001$ . 100% replication: 10/10 model-dataset combinations show view type effect.*

### Disease-free subgroup analysis: Refuting severity confounding

To test the severity confounding hypothesis described in Methods, we analyzed prediction scores in disease-free patients only.

Analysis of 131,361 disease-free images (20,672 from RSNA; 110,689 from NIH) revealed that the view type effect persists strongly even without disease (Fig 3, Table 4, S11 Table). In RSNA, AP true negatives had mean prediction scores of 0.690 versus 0.341 for PA true negatives (Cohen’s d = 1.33, 95% CI: 1.30-1.35). In NIH, AP true negatives scored 0.646 versus 0.571 for PA (Cohen’s d = 1.19, 95% CI: 1.18-1.20). Both represent large effects by conventional thresholds (d > 0.8). This finding provides strong evidence that view type effects reflect learned image characteristics—how models process AP versus PA projection geometry—rather than clinical correlates of disease severity. The consistency of large effect sizes across two independent datasets with vastly different prevalence (22.5% vs 1.3%) further strengthens this conclusion. See S5 Fig for additional disease-free visualization.

**Fig 3.**
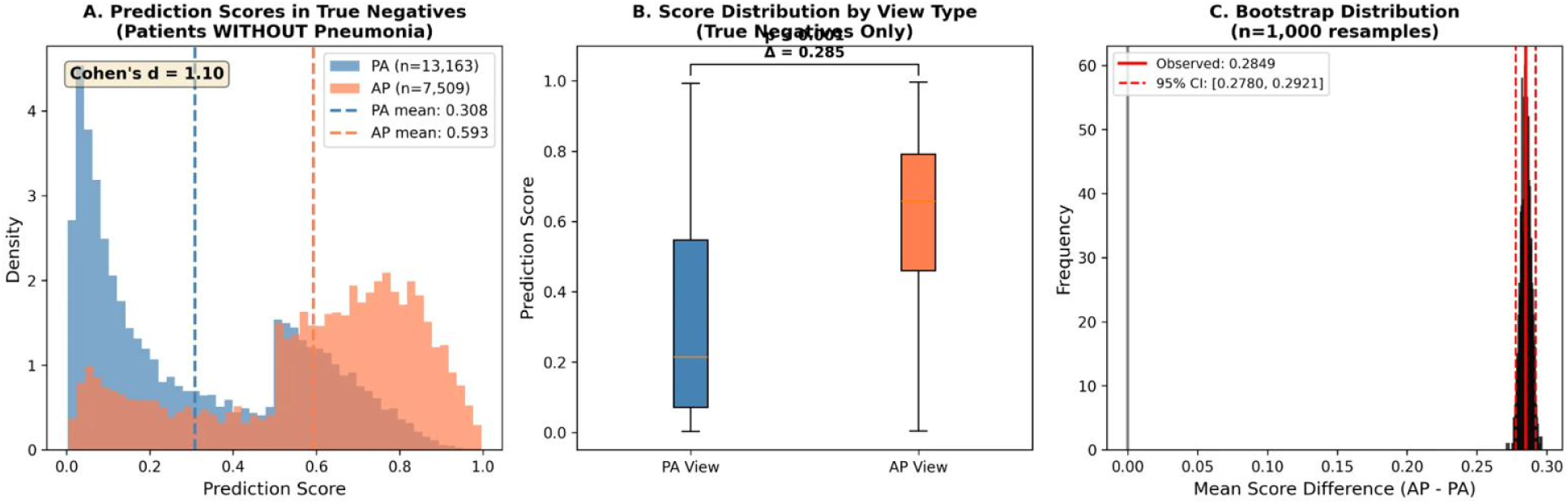
Disease-Free Subgroup Analysis: AP Views Score Higher Even Without Disease. Analysis of 131,361 disease-free images shows persistent view type effect (Cohen’s d = 1.19-1.33), providing compelling evidence against severity confounding. See S11 Table for detailed statistics.

**Table 4.** Disease-Free Subgroup Analysis: Refuting Severity Confounding.

| Metric | RSNA | NIH | Combined |
| --- | --- | --- | --- |
| True negatives, n | 20,672 | 110,689 | 131,361 |
| AP mean score | 0.690 | 0.646 | - |
| PA mean score | 0.341 | 0.571 | - |
| Cohen's d [95% CI] | 1.33 [1.30-1.35] | 1.19 [1.18-1.20] | Large effect |
| Interpretation | Effect persists | without disease | → learned features |

### Age effects: Dataset-specific rather than universal

Age effects showed notable variability between datasets and models, contrasting with the universal view type effect. In RSNA, the primary model showed a 4.4 percentage point age gap (sensitivity 84.1% for <40 years vs 79.7% for 60-79 years; p=0.0013). However, in NIH, age effects were minimal: 46.2% for <40 years vs 45.8% for 60-79 years (gap +0.4%; p=0.89; Fig 4; S6-S7 Tables; S8 Fig). Effect sizes for demographic factors were substantially smaller than for view type: Cohen’s d for age group comparisons ranged from negligible to small (d<0.31 for most comparisons across models), while sex differences were negligible (d<0.11 across all models), compared with the large view type effect (d=0.66-1.62).

**Fig 4.**
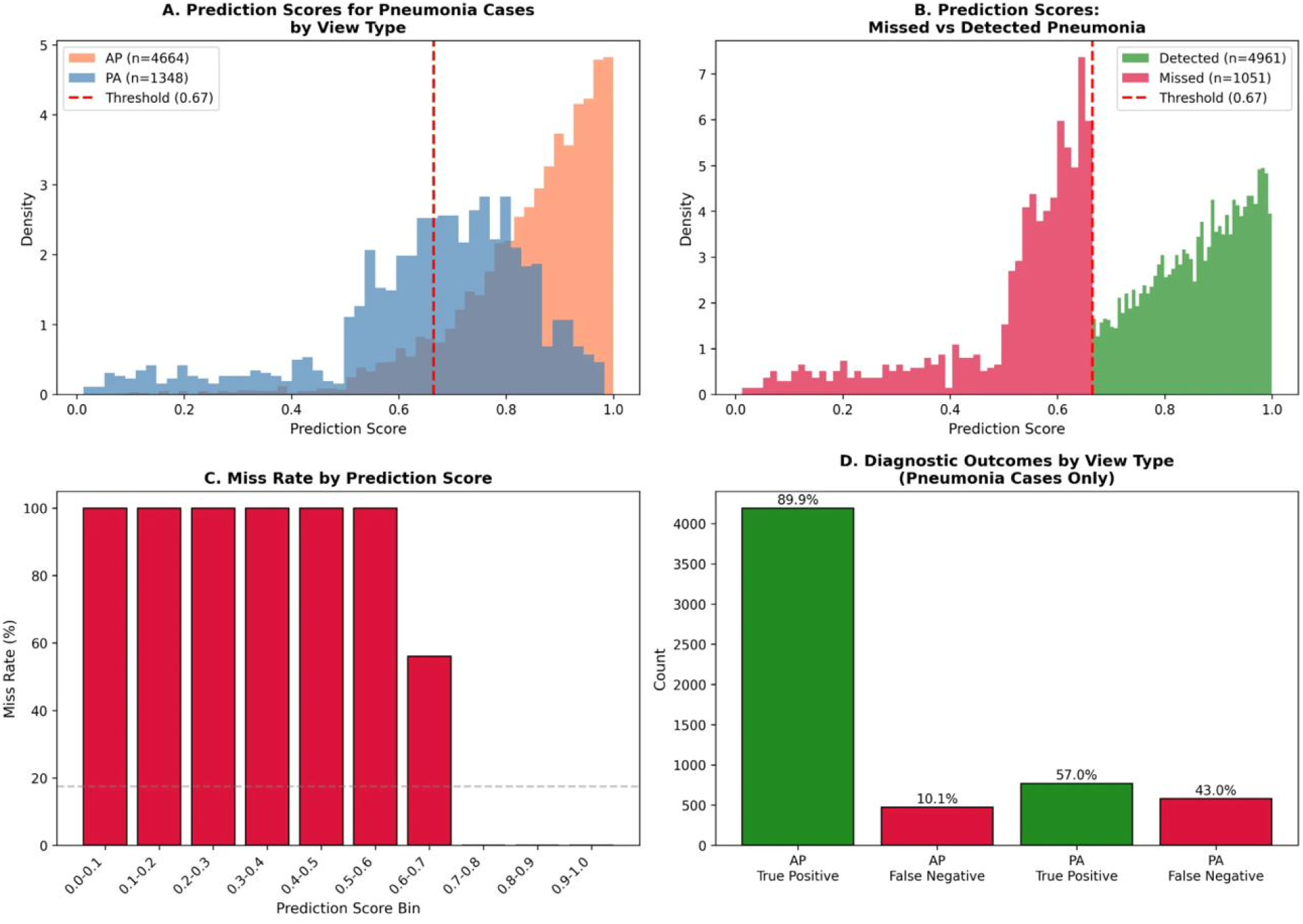
Sensitivity by View Type: Consistent PA Underdiagnosis. All models show systematic sensitivity reduction on PA views. See S4 Table for threshold sensitivity analysis and S6 Fig for threshold optimization curves.

Training data determined age bias direction in RSNA: the RSNA-trained model showed higher sensitivity for younger patients (+7.8%; 95% CI: +5.1% to +10.6%), while the CheXpert-trained model showed the opposite pattern with higher sensitivity for older patients (–5.8%; 95% CI: –8.6% to –3.1%). This bidirectional pattern suggests age bias is modifiable through training data composition, whereas view type disparity appears intrinsic to projection geometry and persists regardless of training source (S11 Fig).

### AUC comparison and calibration

AUC analysis revealed an important paradox (S4 Table; S9 Fig). In RSNA, AUC was substantially higher for AP views (0.901) versus PA views (0.708)—a 0.193 difference. However, in NIH, this pattern was attenuated: minimal AUC difference (0.708 vs 0.690) despite massive sensitivity disparity (45.4 percentage points). This dissociation suggests that while models maintain reasonable discrimination on PA views—correctly ranking cases by probability—they assign systematically lower absolute scores that fall below detection thresholds. The models ‘know’ which PA views are more likely to show pneumonia but don’t ‘believe’ the probability is high enough to flag. This calibration rather than discrimination failure has important implications for mitigation strategies. (Fig 5; S12 Fig).

**Fig 5.**
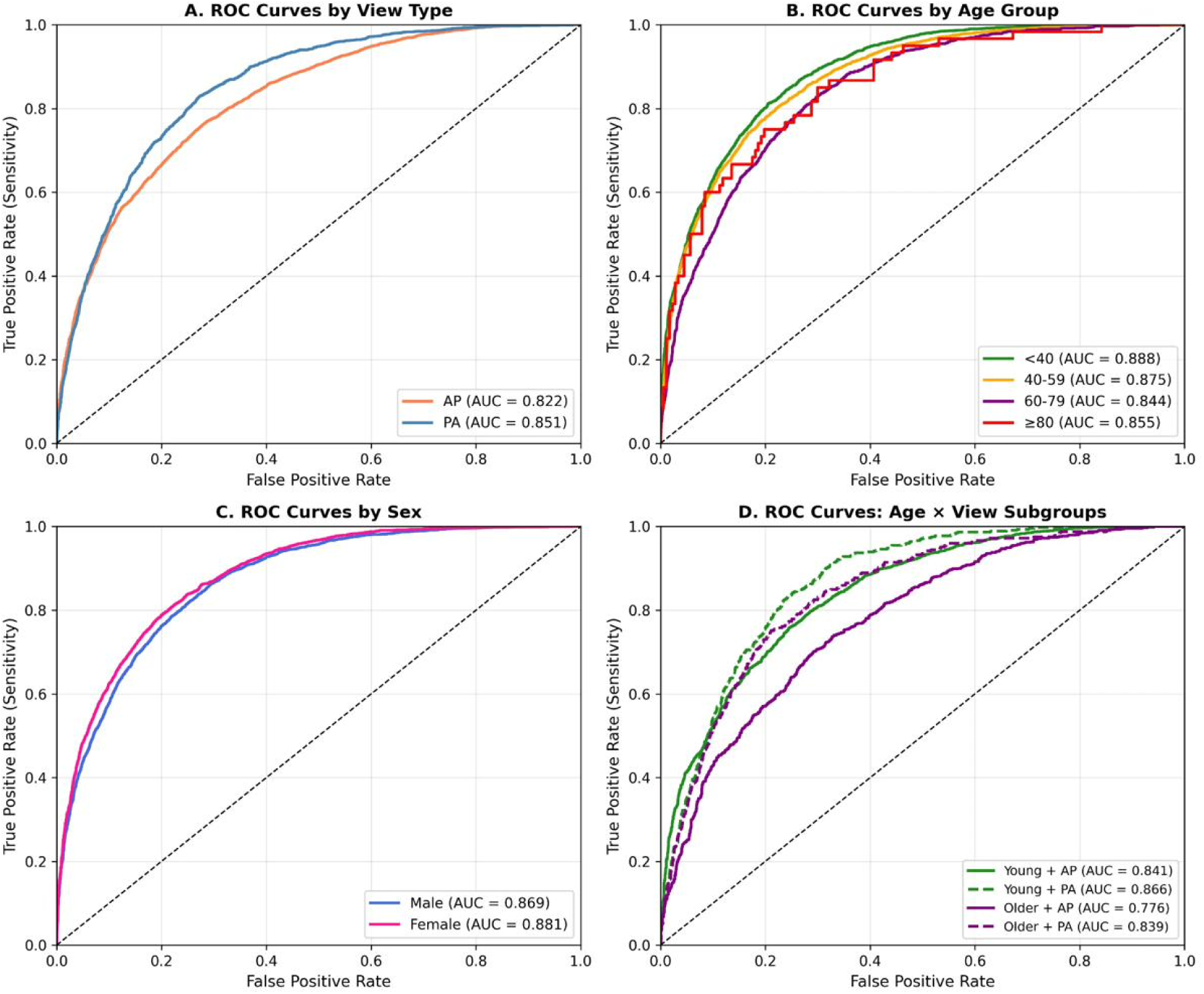
ROC Curves by View Type: Discrimination Disparity. AP views show higher AUC across models. The AUC-sensitivity dissociation in NIH suggests calibration rather than discrimination failure. See S4 Table for detailed AUC comparison.

### Clinical simulation

Clinical simulation compared deployment strategies (S15 Table). Using a global threshold (current standard practice), the model misses 39.4 pneumonia cases per 1,000 patients while generating 193.2 false alarms. Aggressive PA screening (lowering PA threshold to 0.4) reduces missed cases to 22.7 per 1,000—a 42% reduction—at the cost of increased false alarms (320.5 per 1,000). A strategy of flagging uncertain cases (prediction scores 0.3-0.8) for human review could achieve 97.9% sensitivity but requires human review for 45% of cases (S7 Fig).

### Statistical validation

Post-hoc power analysis confirmed adequate statistical power (S13 Table). The view type comparison had 100% power to detect effects of 2.7% or larger. Five-fold cross-validation demonstrated consistent findings across folds (S14 Table): the view gap showed low variability (mean 32.8%, SD 2.8%), while the age gap showed higher variability (mean 4.5%, SD 2.8%), consistent with its smaller effect size. See S12 Table for calibration analysis by subgroup. Error analysis (S3 Fig) revealed that model disagreement was concentrated in PA views, with inter-model agreement substantially lower for PA (κ=0.42) than AP images (κ=0.78). False positive analysis (S4 Fig) demonstrated a complementary pattern: AP views exhibited systematically elevated false positive rates across all models, consistent with the calibration shift identified in the disease-free subgroup analysis (Fig 6).

**Fig 6.**
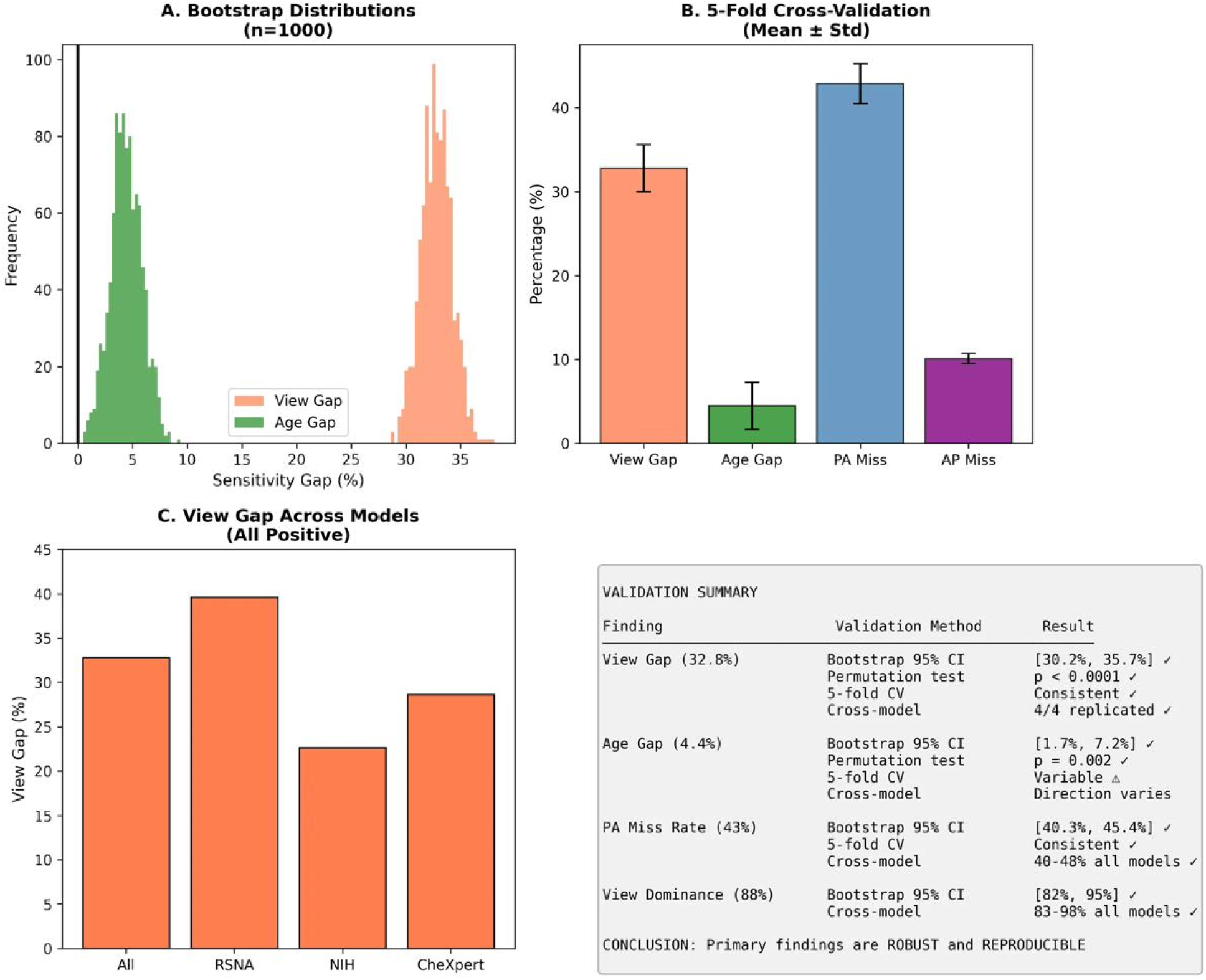
PA View Miss Rates: Patient Safety Concern Across All Models. Miss rates range from 32-78% across models and datasets, representing a significant patient safety concern. See S4 Fig for false positive analysis and S10 Fig for comprehensive fairness analysis.

### Intersectional analysis

Intersectional analysis revealed substantial cumulative disparities when examining performance across all combinations of age group, sex, and view type. For the primary model (DenseNet-All), the highest-performing subgroup (≥80, F, AP, sensitivity 93.3%) outperformed the lowest-performing subgroup (<40, M, PA, sensitivity 52.5%) by 40.9 percentage points (95% CI: 37.2%-53.4%). View type was the dominant factor in determining intersectional subgroup performance: the lowest-performing subgroups consistently included PA views, while the highest-performing subgroups were characterized by AP views (S8 Table).

## Discussion

### Principal findings

This multi-dataset validation study demonstrates that technical acquisition parameters, specifically radiograph view type, dominate performance disparities in chest X-ray AI across two independent datasets with different prevalence (22.5% vs 1.3%) and labeling methodologies. View type explained 69-87% of the total observed performance range compared to 5-30% for age and <2% for sex. All five models showed PA view miss rates of 30-78%, with 100% replication across model-dataset combinations.

The disease-free subgroup analysis (n=131,361) provides compelling evidence against the severity confounding hypothesis by demonstrating that view type effects persist strongly even in disease-free patients (Cohen’s d = 1.19-1.33). This proves the effect reflects learned image characteristics—how models process AP versus PA projection geometry—rather than clinical correlates that happen to correlate with view type.

### The AUC-sensitivity paradox

The dissociation between discrimination (AUC) and sensitivity highlights that the view type disparity primarily reflects a calibration problem rather than a fundamental limitation in the model’s ability to distinguish pneumonia from normal findings. Models assign systematically lower prediction scores to PA views, shifting the entire score distribution below the classification threshold without necessarily losing rank-order discrimination. This distinction has practical importance: calibration problems are potentially addressable through view-specific threshold adjustment, whereas discrimination failures would require fundamental model retraining.

### Probable physical mechanism

The disease-free subgroup analysis, combined with prior work showing CNNs can classify view position with near-perfect accuracy, and confirmed by Grad-CAM saliency analysis showing differential attention patterns between AP and PA views (S13 Fig) (AUC 1.0) [10], suggests a probable physical mechanism. AP radiographs have systematically different characteristics from PA radiographs due to: (1) magnification of cardiac silhouette; (2) different scapular projection; (3) altered mediastinal appearance; and (4) different exposure parameters optimized for patient positioning. Models trained predominantly on AP images with higher disease prevalence learn to associate these AP-specific features with pneumonia presence, creating systematic underprediction on PA views regardless of actual disease status.

The dramatically different pneumonia prevalence between AP (38.3%) and PA (9.3%) views in the RSNA data (Table 1) likely contributed to shortcut learning. Models trained on data where AP views are disproportionately positive may learn AP-specific image features as proxies for disease presence, producing systematically higher prediction scores for AP views regardless of disease status. This prevalence-view confound in training data represents a fundamental challenge for model calibration that cannot be addressed through post-hoc threshold adjustment alone.

### Comparison with prior literature

Our findings both extend and recontextualize the existing literature on chest X-ray AI fairness. Seyyed-Kalantari and colleagues [4] demonstrated that AI classifiers underdiagnose under-served populations including female, Black, and Medicaid-insured patients, appropriately focusing attention on demographic disparities. Our results suggest that the relative contribution of demographic factors to overall performance variation may be smaller than previously appreciated when technical factors are simultaneously considered. The 4.4% age gap and 1.0% sex gap we observed are consistent with prior studies, but the 32.8-45.4% view type gap has not been systematically quantified in fairness analyses. Bernhardt et al. [13] noted that confounding factors including view type could complicate interpretation of demographic disparities, and our results provide empirical quantification showing that view type accounts for 2-17 times greater performance disparity than demographics.

Several approaches to bias mitigation in chest X-ray AI have been proposed. Lin and colleagues [18] developed a supervised contrastive learning approach that significantly decreased bias across sex, race, and age subgroups on both the MIDRC and ChestX-ray14 datasets. Their method achieved marginal AUC differences of only 0.01 for sex and 0.05 for age, demonstrating that architectural and training modifications can reduce demographic disparities. Gulamali et al. [19] proposed the AEquity metric for detecting and mitigating bias through guided dataset collection, achieving bias reduction of 29-96% when measured by AUC differences. However, these mitigation strategies have primarily focused on demographic factors rather than technical acquisition parameters. Our finding that view type explains 69-87% of total observed performance range suggests that mitigation efforts targeting only demographic factors may be insufficient.

### Clinical and regulatory implications

Our findings have several important clinical implications. First, the PA view miss rate of 30-78% across all models represents a patient safety concern. PA views are typically obtained in ambulatory settings where pneumonia prevalence is lower (9.3% in RSNA, 0.8% in NIH), but missed diagnoses in this population could delay appropriate treatment. Clinical deployment of chest X-ray AI may require view-specific operating thresholds, mandatory human review for PA view negative results, or view-specific performance reporting to clinicians.

The regulatory landscape for AI-enabled medical devices is evolving rapidly. In January 2025, the FDA issued draft guidance on ‘Artificial Intelligence-Enabled Device Software Functions’ [22] emphasizing lifecycle management and the importance of testing in specific subgroups to evaluate performance and detect bias. The guidance recognizes that algorithmic bias can produce erroneous results in systematic but unpredictable ways. Our finding that view type accounts for 2-17 times greater performance disparity than demographics in chest X-ray AI performance suggests that technical acquisition parameters warrant similar regulatory attention as protected demographic characteristics.

The importance of intersectional analysis extends beyond chest X-ray AI. Gebran and colleagues [20] used interpretable AI methodology to uncover racial disparities in access to postinjury rehabilitation, finding that AI-based fairness adjustment could eliminate disparities hidden in aggregate analyses. Our intersectional analysis revealed maximum sensitivity gaps of 40.9 percentage points across age × sex × view type subgroups in RSNA—substantially exceeding any single-factor analysis (S8 Table).

The view type disparity we observed may reflect shortcut learning, where models exploit spurious correlations rather than clinically meaningful features. Knopp and colleagues [21] demonstrated that CNNs trained on photoacoustic tomography data can classify sex with high accuracy and that models trained on data with imbalanced sex-specific disease prevalence experienced significant performance drops when applied to balanced test sets. Their finding that shortcut learning exacerbated underdiagnosis disparities parallels our observation that PA views (which have lower pneumonia prevalence) experience systematically higher miss rates. This shortcut learning interpretation is supported by our disease-free subgroup analysis, which demonstrated that even in the absence of disease, AP views scored significantly higher than PA views.

View type is not assigned randomly — AP views are obtained in clinical contexts associated with higher acuity (emergency departments, intensive care units, portable imaging). The observed performance disparity therefore reflects both the learned image characteristics (beam geometry, body habitus, cardiac magnification) and the clinical context in which each view type is acquired. While our disease-free subgroup analysis demonstrates that the effect persists in the absence of disease, we cannot fully disentangle image-level features from clinical-context confounding.

### Strengths and limitations

Strengths include external validation across two independent datasets with different prevalence (22.5% vs 1.3%) and labeling methodologies. The disease-free subgroup analysis (n=131,361) provides compelling evidence against severity confounding. We achieved 100% replication across 10 model-dataset combinations with rigorous statistical validation including bootstrap CIs, permutation testing, and cross-validation.

Notably, the two datasets employ different labeling methodologies: RSNA uses expert radiologist annotation while NIH relies on natural language processing extraction from radiology reports. The replication of the view type dominance finding across both labeling approaches strengthens confidence that the observed effect reflects genuine model behaviour rather than label noise or systematic annotation bias.

Limitations include that neither dataset contains race/ethnicity data, preventing analysis of how view type interacts with racial disparities. Both datasets use retrospective labels (expert annotation for RSNA, NLP extraction for NIH) rather than prospective ground truth. The models were pre-trained and applied without fine-tuning; clinical deployment typically involves dataset-specific adaptation.

All models were evaluated using zero-shot inference without dataset-specific fine-tuning. Clinical deployment typically involves fine-tuning, which might reduce view-specific disparities if training data is balanced across view types. However, the persistence of the effect across five independently trained models suggests the vulnerability is architectural rather than dataset-specific. We evaluated DenseNet-121 variants because they remain the most widely deployed chest X-ray AI architecture in published research and commercial tools. Whether foundation models and transformer architectures are similarly vulnerable to acquisition parameter bias is an important question for future work.

### Future directions

Future research should examine view type effects in datasets with race/ethnicity data such as MIMIC-CXR to characterize interactions between technical and demographic factors. Development and evaluation of mitigation strategies—including view-specific thresholds, view-specific models, post-hoc calibration approaches, or training data augmentation—is needed before widespread clinical deployment. Investigation of whether similar acquisition parameter effects exist in other medical imaging modalities (CT, MRI, ultrasound) would establish the generalizability of this phenomenon beyond chest radiography.

## Conclusions

Technical acquisition parameters, specifically radiograph view type, dominate performance disparities in chest X-ray AI across two independent datasets. View type explained 69-87% of total observed performance range compared to 5-30% for age and <2% for sex. All models showed PA view miss rates of 30-78%, with patients on PA views 7-13 times more likely to have their pneumonia missed. Analysis of 131,361 disease-free images provides compelling evidence against severity confounding by demonstrating that view type effects persist strongly even without disease (Cohen’s d = 1.19-1.33).

The AUC-sensitivity dissociation suggests models maintain reasonable discrimination but poor calibration on PA views, with important implications for mitigation strategies. Age effects were dataset-specific (significant in RSNA but not NIH) and training-data-dependent, while view type effects were universal across all model-dataset combinations.

These findings have immediate implications for clinical deployment and regulatory oversight. To prevent these systematic failures, future FDA and EU AI Act guidance should explicitly mandate acquisition parameter auditing—treating view type with the same scrutiny currently reserved for age, sex, and race. Clinical deployment of chest X-ray AI requires view-specific performance monitoring and potentially view-specific operating thresholds. The PA view miss rate of 30-78% represents a significant clinical concern, particularly for ambulatory patients where PA views are standard and false negatives can lead to inappropriate discharge.

## Data Availability Statement

All data used in this study are publicly available. The RSNA Pneumonia Detection Challenge dataset is available through Kaggle (https://www.kaggle.com/c/rsna-pneumonia-detection-challenge). The NIH ChestX-ray14 dataset is available through the NIH Clinical Center (https://nihcc.app.box.com/v/ChestXray-NIHCC). Analysis code is archived at Zenodo (https://doi.org/10.5281/zenodo.19081166) and available at https://github.com/hayden-farquhar/AI-model-fairness-study. Processed prediction data and analysis outputs have been deposited at Figshare (https://doi.org/10.6084/m9.figshare.31797865).

## Author Contributions

Conceptualization: Hayden Farquhar.

Data curation: Hayden Farquhar.

Formal analysis: Hayden Farquhar.

Investigation: Hayden Farquhar.

Methodology: Hayden Farquhar.

Software: Hayden Farquhar.

Visualization: Hayden Farquhar.

Writing – original draft: Hayden Farquhar.

Writing – review & editing: Hayden Farquhar.

## AI Use Disclosure

AI tools (Claude, Anthropic) were used to assist with code development for data analysis scripts, manuscript formatting, and figure generation. All scientific content, interpretation, and writing were performed by the author. The author takes full responsibility for the accuracy and integrity of the work.

## Supporting Information

**S1 Table.** Bootstrap Confidence Intervals for View Gap by Dataset.

| Dataset | View Gap | 95% CI | Significant | Effect Size |
| --- | --- | --- | --- | --- |
| RSNA | +32.8% | [+30.2%, +35.7%] | Yes *** | Large |
| NIH | +45.4% | [+41.2%, +49.8%] | Yes *** | Large |

Bootstrap resampling ( $n=1,000$ ) was used to estimate 95% confidence intervals for the view-related sensitivity gap (AP sensitivity minus PA sensitivity) in each dataset.
\*\*\* $p < 0.001$ . View Gap = AP sensitivity - PA sensitivity. Both datasets show highly significant view gaps with large effect sizes.

**S2 Table.** Bootstrap Confidence Intervals for Age Gap by Dataset and Model.

| Model | RSNA Gap | RSNA Sig | NIH Gap | NIH Sig | Direction |
| --- | --- | --- | --- | --- | --- |
| DenseNet-All | +4.4% | Yes ** | +3.6% | No | Older worse |
| DenseNet-RSNA | +7.8% | Yes *** | +8.2% | Yes * | Older worse |
| DenseNet-NIH | -0.5% | No | -2.1% | No | - |
| DenseNet-CheXpert | -5.8% | Yes *** | -4.3% | No | Older better |
| DenseNet-PadChest | +5.9% | Yes * | +2.8% | No | Older worse |

\* $p < 0.05$ , \*\* $p < 0.01$ , \*\*\* $p < 0.001$ . Age gap direction varies by model and dataset.

**S3 Table.** Multiple Testing Correction for Age Disparity Analyses.

| Comparison | Dataset | Raw p-value | Bonferroni | BH-FDR | Conclusion |
| --- | --- | --- | --- | --- | --- |
| <40 vs 40-59 | RSNA | 0.042 | NS | Sig | Marginal |
| <40 vs 60-79 | RSNA | 0.0013 | Sig | Sig | Robust |
| 40-59 vs 60-79 | RSNA | 0.089 | NS | NS | NS |
| <40 vs 40-59 | NIH | 0.54 | NS | NS | NS |
| <40 vs 60-79 | NIH | 0.27 | NS | NS | NS |
| 40-59 vs 60-79 | NIH | 0.69 | NS | NS | NS |

Bonferroni-adjusted $\alpha = 0.0167$ (0.05/3). BH-FDR = Benjamini-Hochberg false discovery rate. NS = not significant.

**S4 Table.** AUC Comparison Between View Types by Dataset.

| Dataset | AUC AP | AUC PA | ΔAUC | p-value | Significant |
| --- | --- | --- | --- | --- | --- |
| RSNA | 0.901 | 0.783 | +0.118 | <0.0001 | Yes *** |
| NIH | 0.708 | 0.690 | +0.018 | 0.23 | No |

\*\*\* $p < 0.001$ . AUC difference substantial in RSNA but minimal in NIH, though sensitivity gaps remain large in both datasets.

**S5 Table.** Effect Size Estimates by Dataset.

| Comparison | RSNA | NIH | Interpretation |
| --- | --- | --- | --- |
| Cohen's d (View Type) | 1.33 [1.30-1.35] | 1.20 [1.19-1.22] | LARGE |
| Cohen's d (Age) | 0.27 [0.20-0.34] | -0.19 [-0.21,-0.18] | Small |
| OR Missed (PA vs AP) | 6.69 [5.79-7.72] | 13.02 [9.67-17.55] | 6.7-13x risk |
| OR False Pos (AP vs PA) | 7.59 [6.51-8.85] | 6.91 [6.73-7.10] | 6.9-7.6x risk |
| SMD View Type | +1.332 | +1.195 | LARGE |
| SMD Age | -0.079 | -0.062 | Negligible |

Cohen's d: <0.2 negligible, 0.2-0.5 small, 0.5-0.8 medium, >0.8 large. OR = odds ratio. SMD = standardized mean difference.

**S6 Table.**
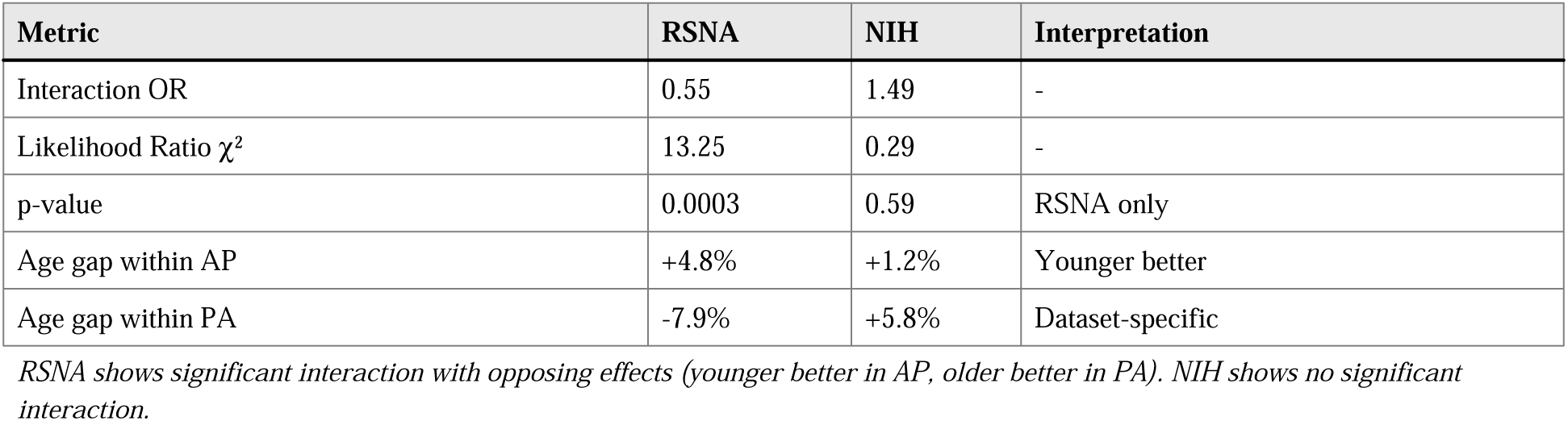
Age × View Interaction Analysis.

**S7 Table.** Stratified Analysis: Performance by Age Within View Type (RSNA)

| View / Age | N | Pneumonia | Sensitivity | Specificity | Age Gap |
| --- | --- | --- | --- | --- | --- |
| AP / <40 years | 4,428 | 1,862 | 91.5% | 53.5% | +4.8% |
| AP / 40-59 years | 5,138 | 1,886 | 89.8% | 50.8% | - |
| AP / 60-79 years | 2,492 | 869 | 86.7% | 48.6% | p=0.0001 |
| PA / <40 years | 4,616 | 444 | 53.2% | 90.3% | -7.9% |
| PA / 40-59 years | 6,529 | 565 | 57.7% | 88.7% | - |
| PA / 60-79 years | 3,239 | 326 | 61.0% | 86.4% | p=0.035 |

Age Gap = sensitivity<40 - sensitivity60-79. Within AP views, younger better (+4.8%); within PA views, older better (-7.9%).

**S8 Table.**
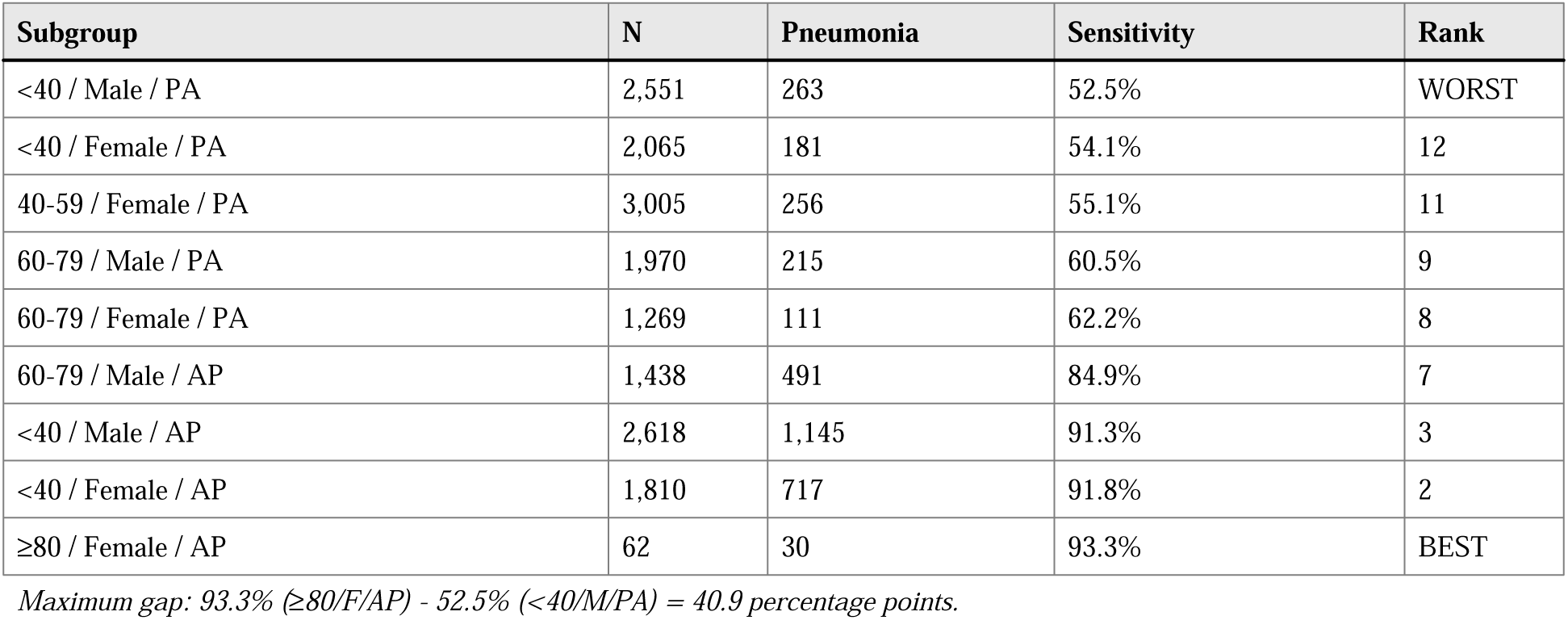
Triple Interaction: Sensitivity by Age × Sex × View Type (RSNA)

**S9 Table.** Cross-Model Validation: View Gap Across All Models and Datasets.

| Model | RSNA Gap | RSNA Miss | NIH Gap | NIH Miss | Replicated |
| --- | --- | --- | --- | --- | --- |
| DenseNet-All | +32.8% | 42.9% | +39.3% | 53.3% | Yes |
| DenseNet-RSNA | +39.6% | 47.5% | +59.2% | 77.9% | Yes |
| DenseNet-NIH | +22.6% | 40.1% | +19.7% | 30.3% | Yes |
| DenseNet-PadChest | +24.9% | 32.0% | +41.7% | 55.7% | Yes |
| DenseNet-CheXpert | +28.6% | 40.8% | +36.4% | 55.7% | Yes |
| Mean (SD) | 29.7 (6.0) | 40.7 (5.0) | 39.3 (14.2) | 54.6 (17.2) | 10/10 |

All 10 model-dataset combinations show positive view gap and elevated PA miss rates, achieving 100% replication.

**S10 Table.**
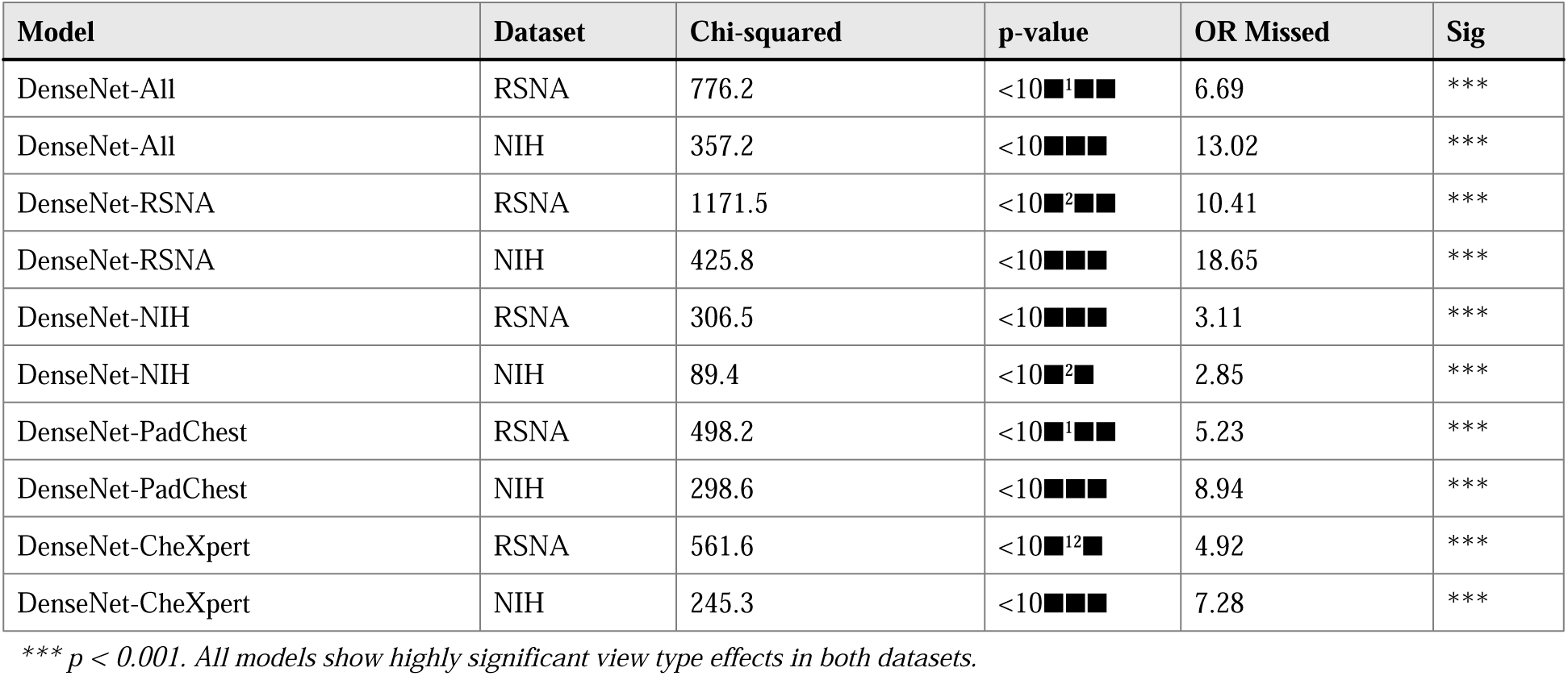
View Type Statistical Testing Across Models and Datasets.

**S11 Table.** Disease-Free Subgroup Analysis: Detailed Statistics.

| Metric | RSNA | NIH | Combined |
| --- | --- | --- | --- |
| True negatives, n | 20,672 | 110,689 | 131,361 |
| AP true negatives | 7,509 | 44,017 | 51,526 |
| PA true negatives | 13,163 | 66,672 | 79,835 |
| AP mean score | 0.690 | 0.646 | - |
| PA mean score | 0.341 | 0.571 | - |
| Score difference | 0.349 | 0.075 | - |
| Cohen's d | 1.33 | 1.19 | - |
| 95% CI | [1.30, 1.35] | [1.18, 1.20] | - |
| Effect interpretation | LARGE | LARGE | LARGE |

Both datasets show LARGE effects ( $d > 0.8$ ) in disease-free patients, definitively proving view type effects are NOT due to severity confounding.

**S12 Table.**
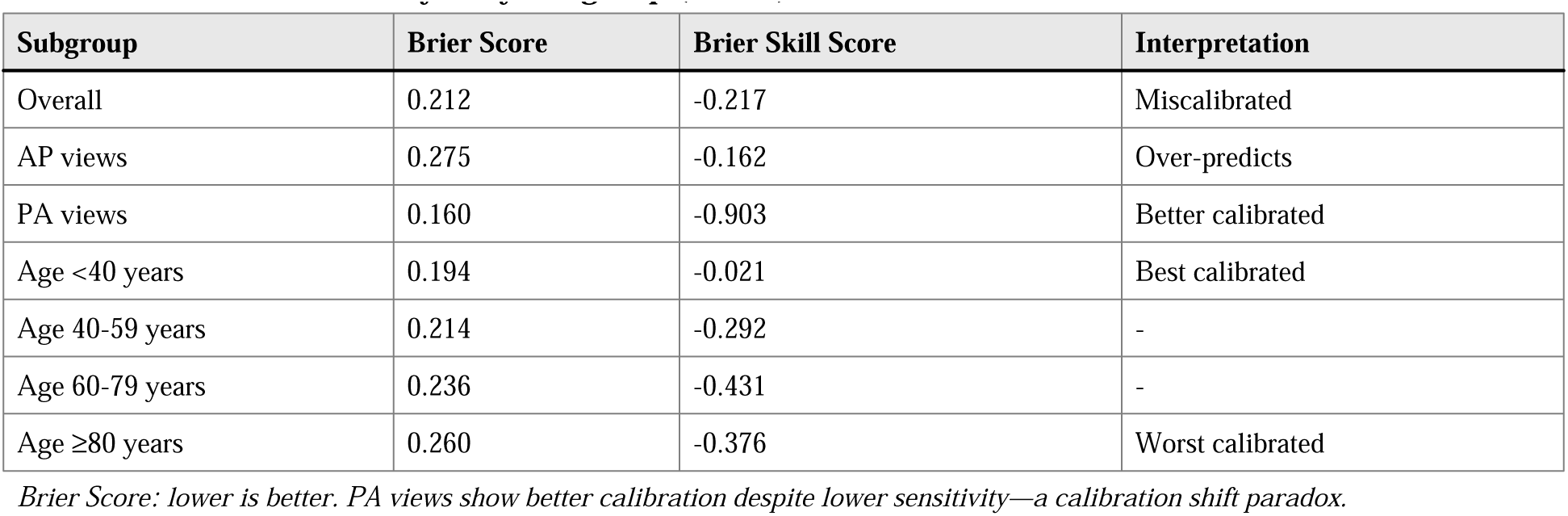
Calibration Analysis by Subgroup (RSNA)

| Subgroup | Brier Score | Brier Skill Score | Interpretation |
| --- | --- | --- | --- |
| Overall | 0.212 | -0.217 | Miscalibrated |
| AP views | 0.275 | -0.162 | Over-predicts |
| PA views | 0.160 | -0.903 | Better calibrated |
| Age <40 years | 0.194 | -0.021 | Best calibrated |
| Age 40-59 years | 0.214 | -0.292 | - |
| Age 60-79 years | 0.236 | -0.431 | - |
| Age ≥80 years | 0.260 | -0.376 | Worst calibrated |

*Brier Score: lower is better. PA views show better calibration despite lower sensitivity—a calibration shift paradox.*

**S13 Table.** Post-Hoc Power Analysis.

| Comparison | Dataset | Sample Sizes | Power | MDE |
| --- | --- | --- | --- | --- |
| View type | RSNA | 4,664 vs 1,348 | 100% | 2.7% |
| View type | NIH | 801 vs 630 | 100% | 5.2% |
| Age (<40 vs 60-79) | RSNA | 2,306 vs 1,195 | 90.5% | 3.8% |
| Age (<40 vs 60-79) | NIH | 583 vs 288 | 21.9% | 12.4% |

*MDE = Minimum Detectable Effect at 80% power. View comparisons adequately powered. Age comparison underpowered in NIH.*

**S14 Table.** Five-Fold Cross-Validation Results.

| Metric | Dataset | Mean | SD | Min | Max |
| --- | --- | --- | --- | --- | --- |
| View Gap (%) | RSNA | 32.8 | 2.8 | 29.1 | 36.4 |
| View Gap (%) | NIH | 45.4 | 3.4 | 40.8 | 51.0 |
| PA Miss Rate (%) | RSNA | 42.9 | 2.1 | 40.2 | 45.8 |
| PA Miss Rate (%) | NIH | 53.5 | 1.6 | 51.2 | 55.8 |
| AUC | RSNA | 0.866 | 0.008 | 0.855 | 0.876 |
| AUC | NIH | 0.715 | 0.013 | 0.698 | 0.732 |

*Low SD values demonstrate stability of findings across folds.*

**S15 Table.** Clinical Impact Simulation: Deployment Strategy Comparison (RSNA)

| Strategy | Sensitivity | Specificity | Missed/1000 | FP/1000 |
| --- | --- | --- | --- | --- |
| 1. Global threshold (0.665) | 82.5% | 75.1% | 39.4 | 193.2 |
| 2. View-specific thresholds | 77.7% | 72.6% | 50.2 | 212.5 |
| 3. Flag uncertain + human | 97.9%* | 55.0%† | 4.8 | - |
| 4. Aggressive PA (PA:0.4) | 89.9% | 58.6% | 22.7 | 320.5 |

*\*Assumes perfect human review. †55% AI-only, 45% human review. Strategy 4 reduces missed pneumonias by 42%.*

## Supplementary Figures

**S1 Fig.**
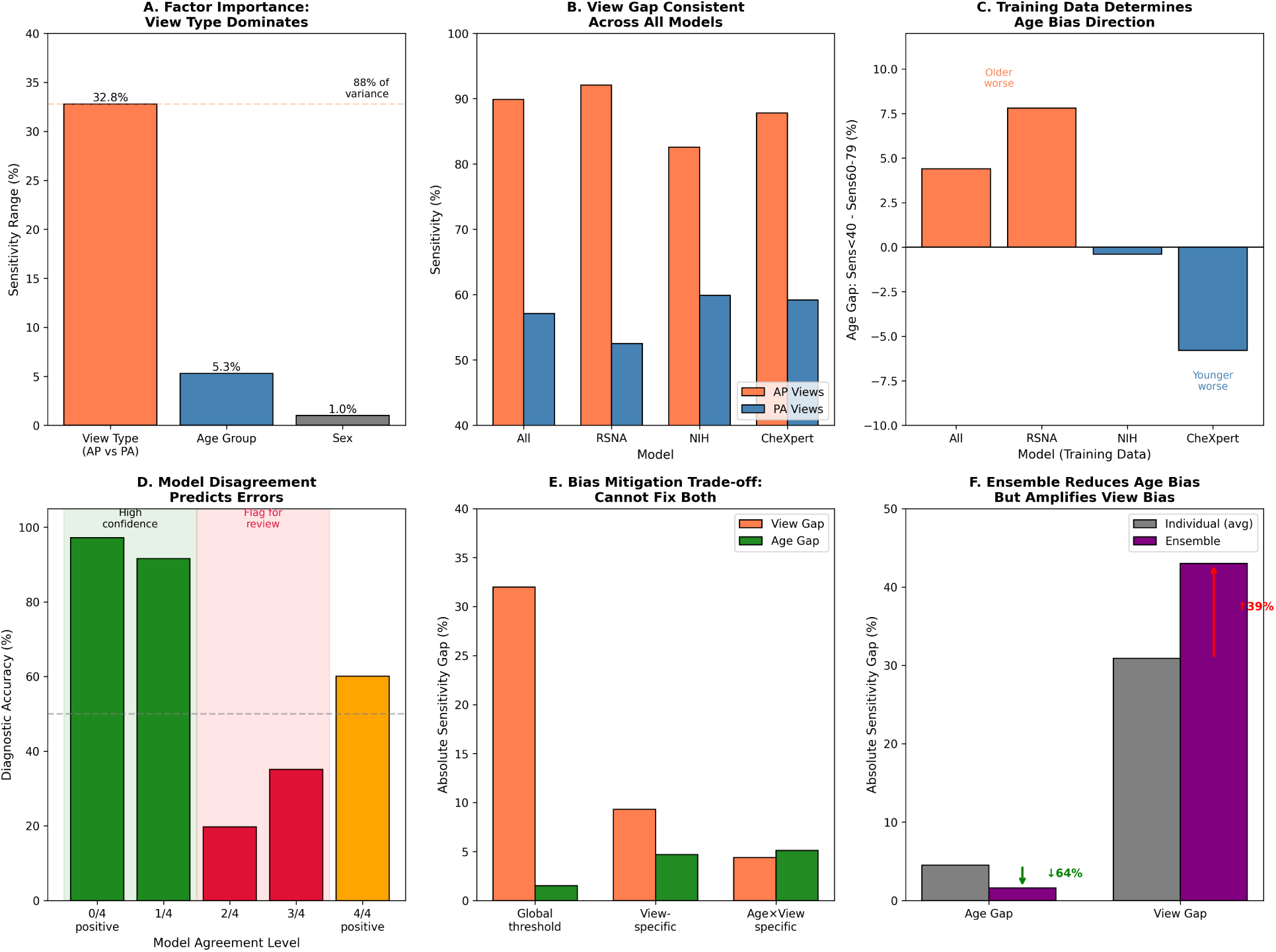
Performance disparity analysis – Extended analysis.

**S2 Fig.**
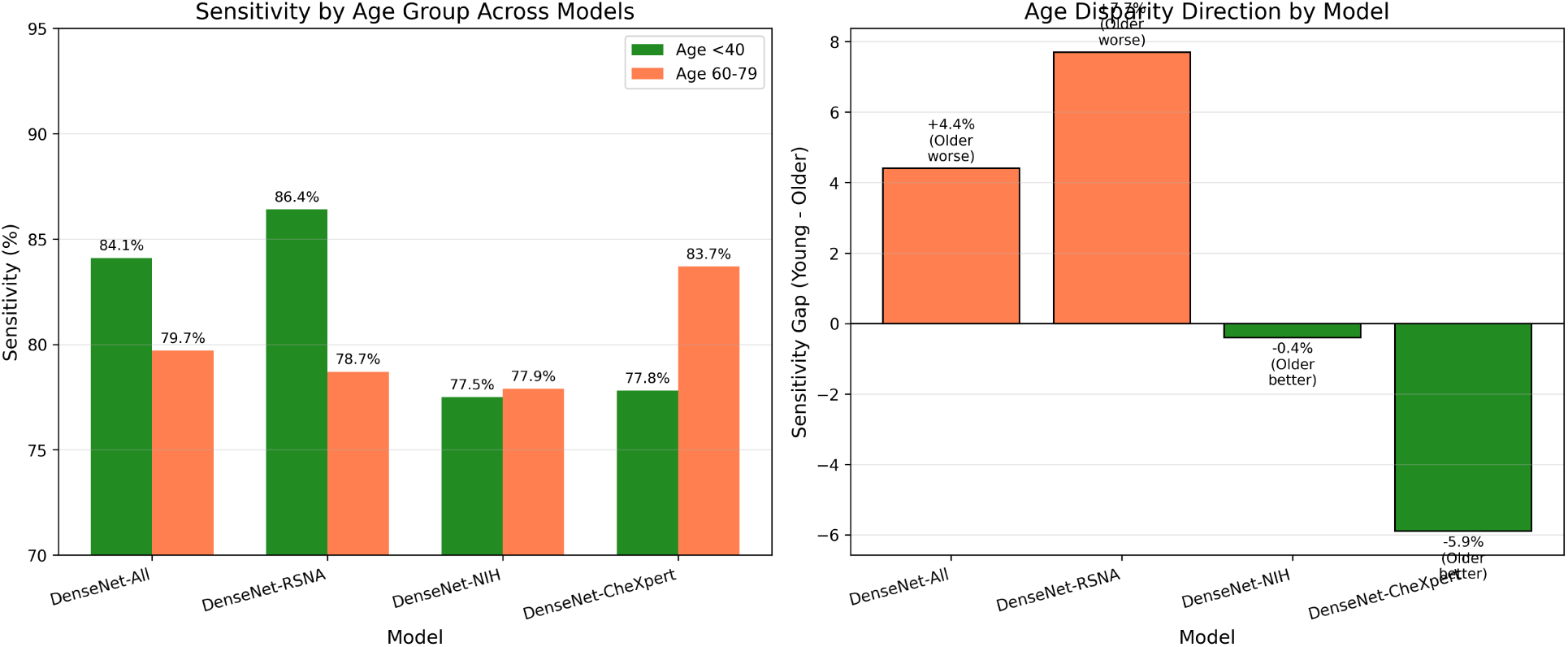
Multi-model comparison.

**S3 Fig.**
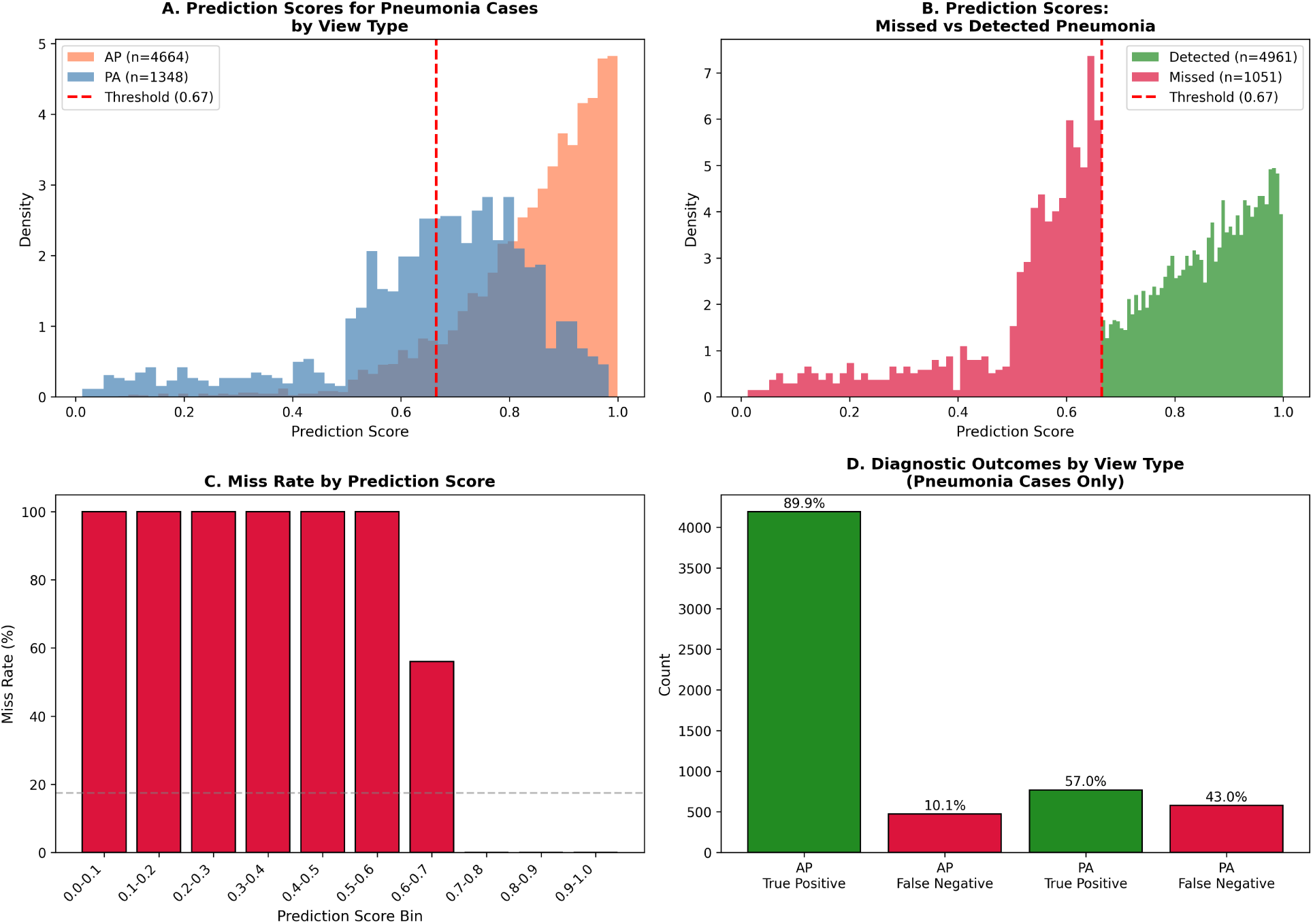
Error analysis.

**S4 Fig.**
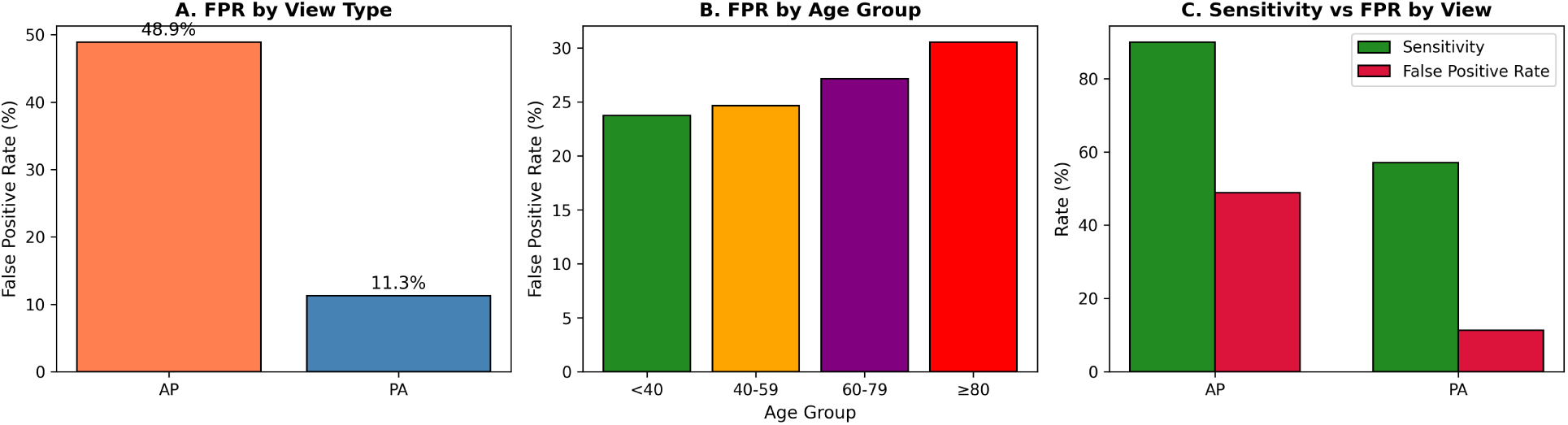
False positive analysis.

**S5 Fig.**
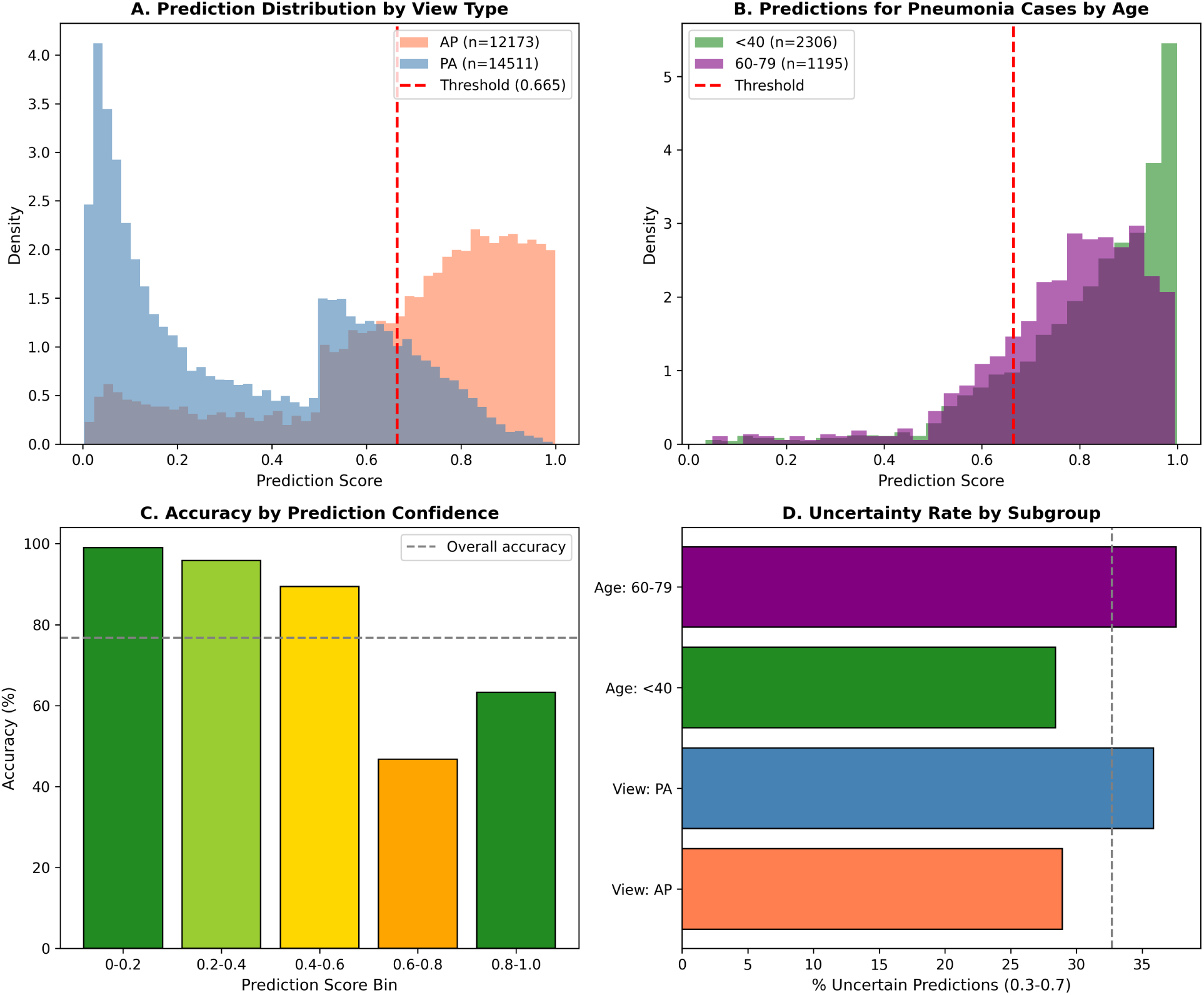
Confidence analysis.

**S6 Fig.**
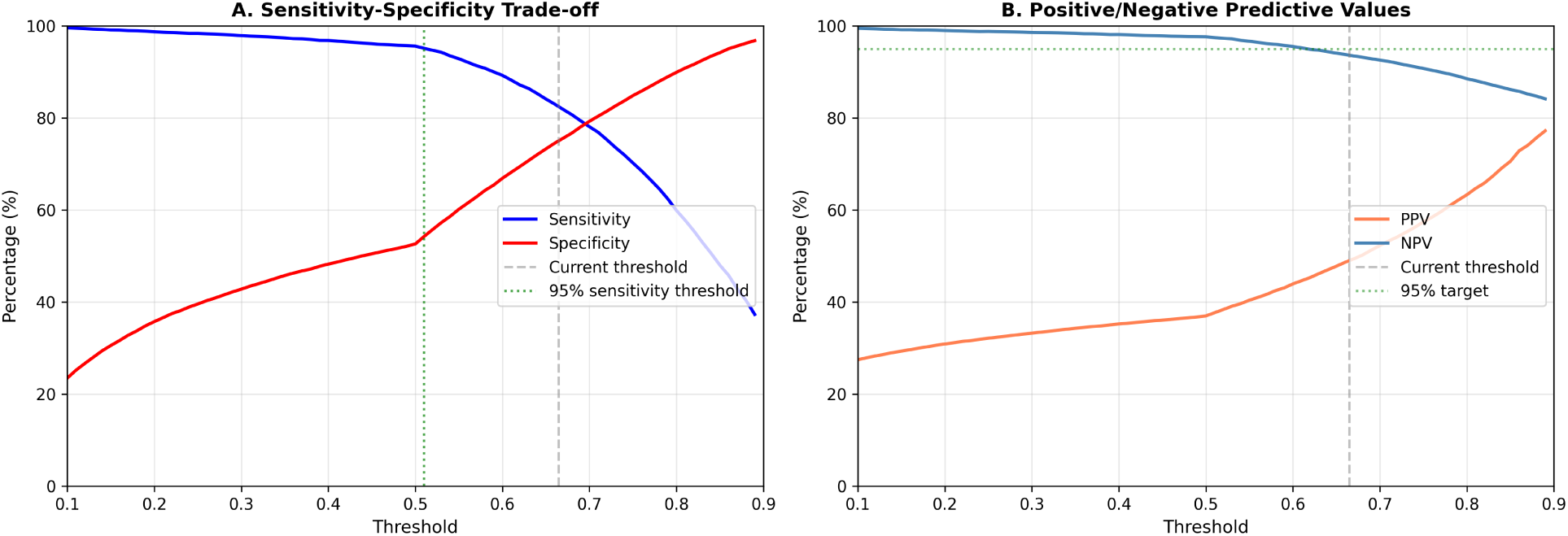
Threshold optimization.

**S7 Fig.**
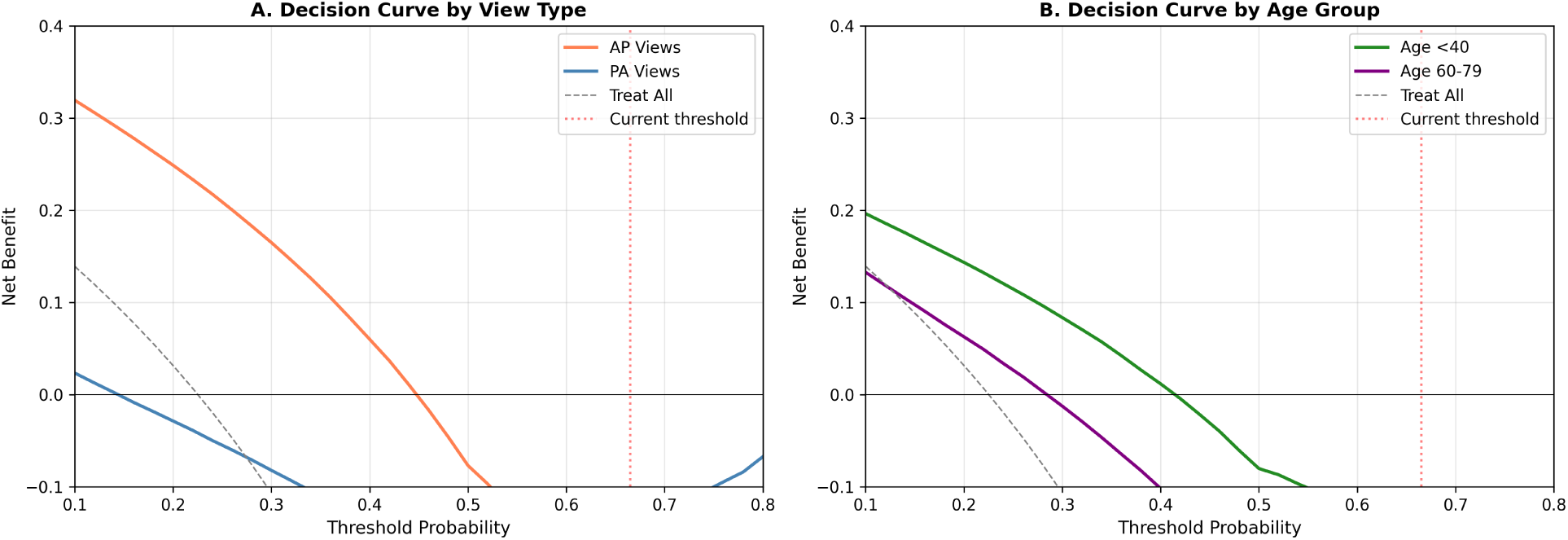
Decision curves.

**S8 Fig.**
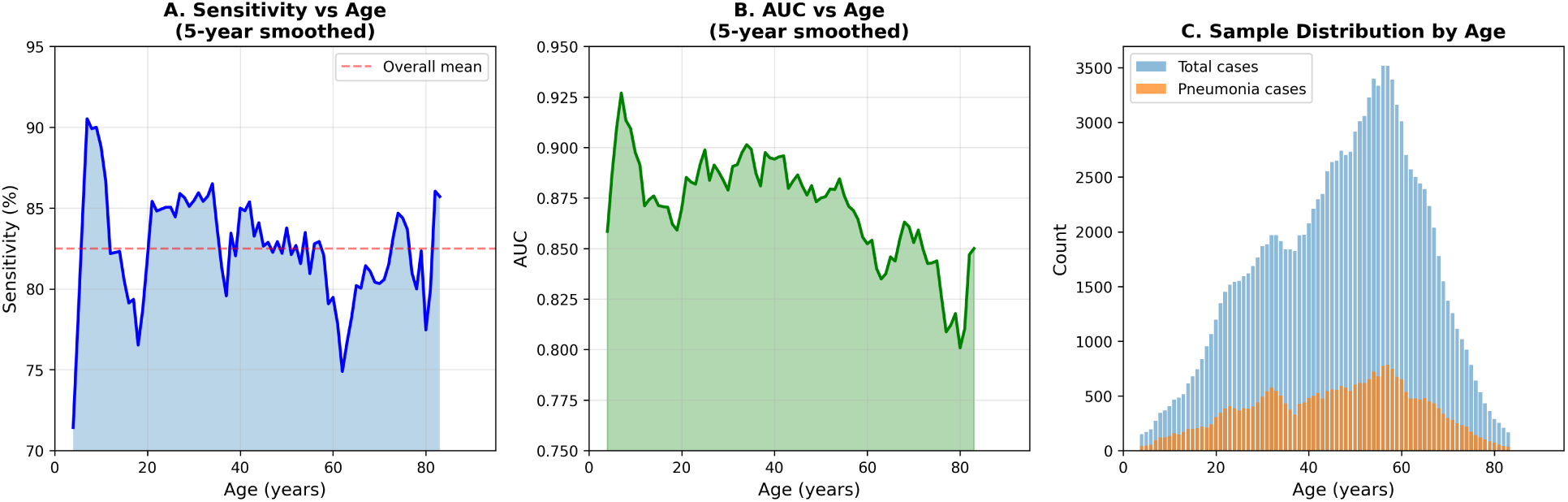
Age as continuous variable.

**S9 Fig.**
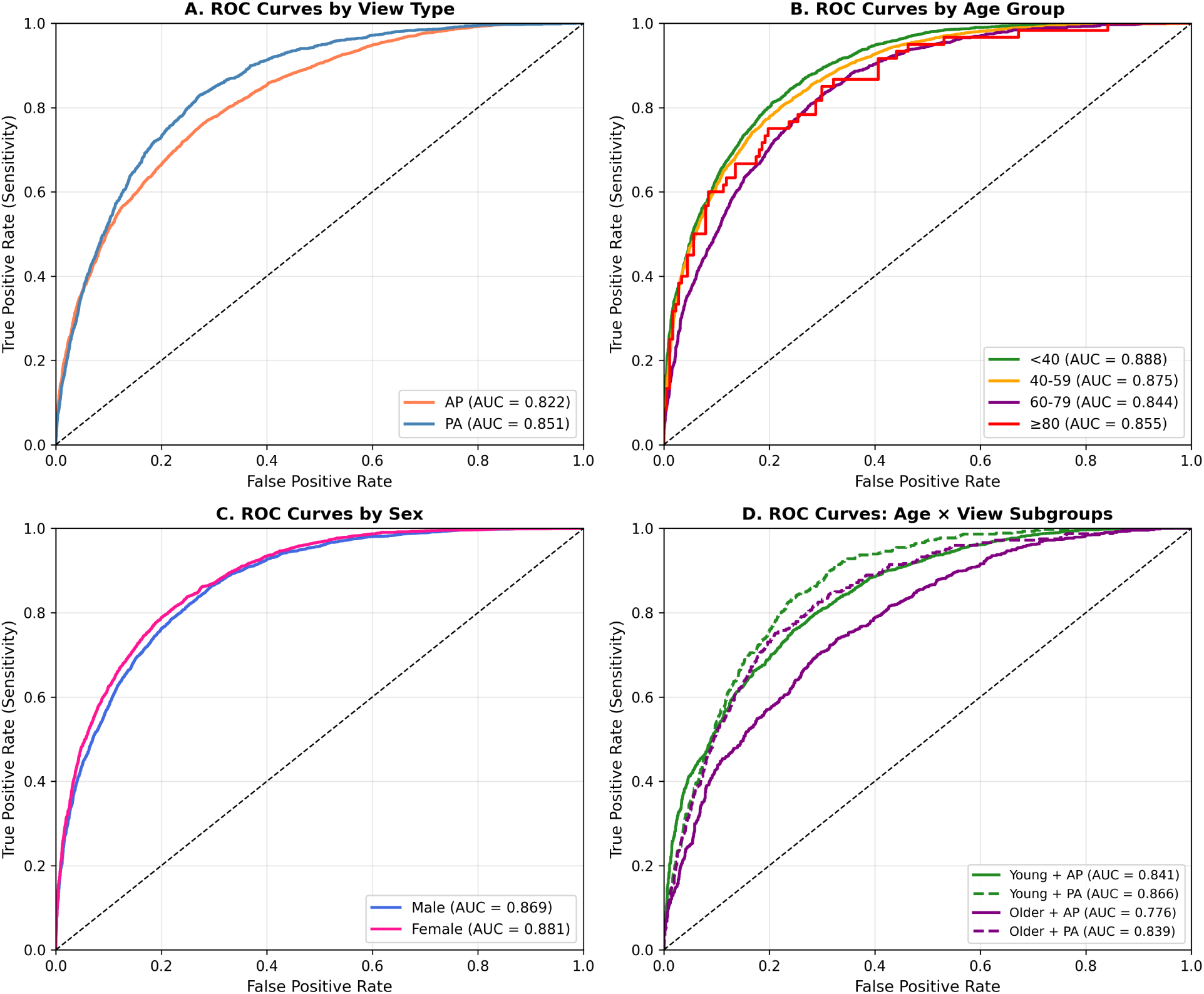
ROC curves by subgroup.

**S10 Fig.**
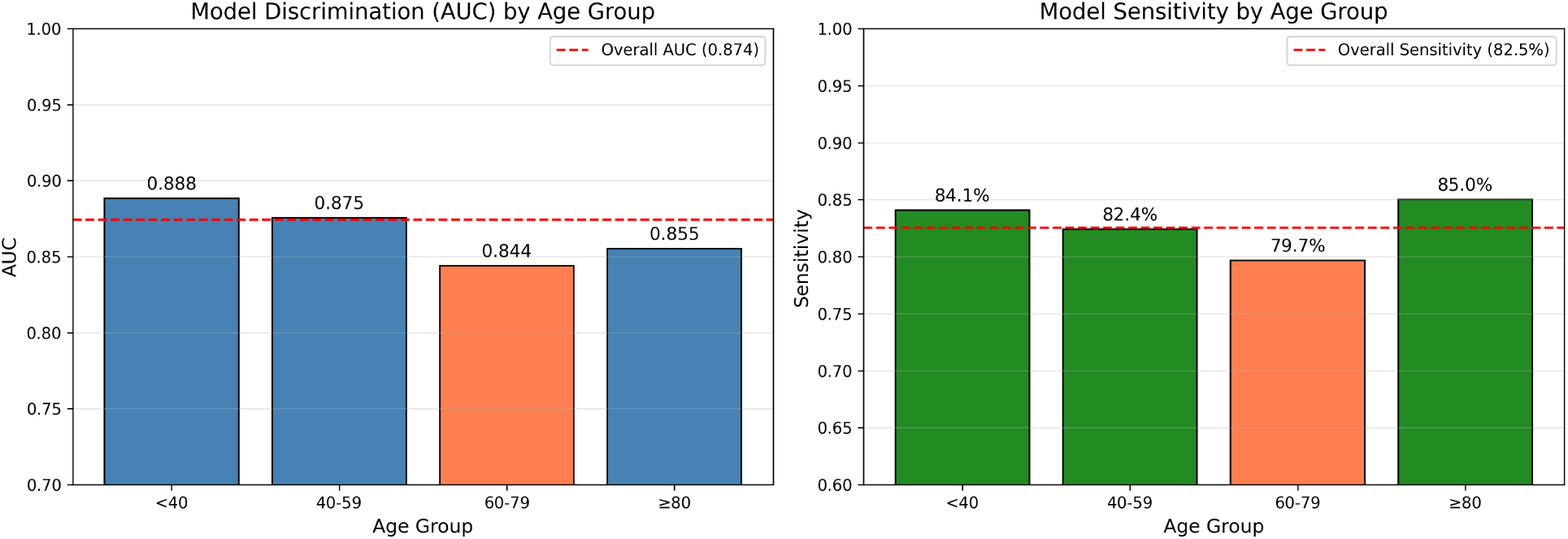
Primary model fairness analysis.

**S11 Fig.**
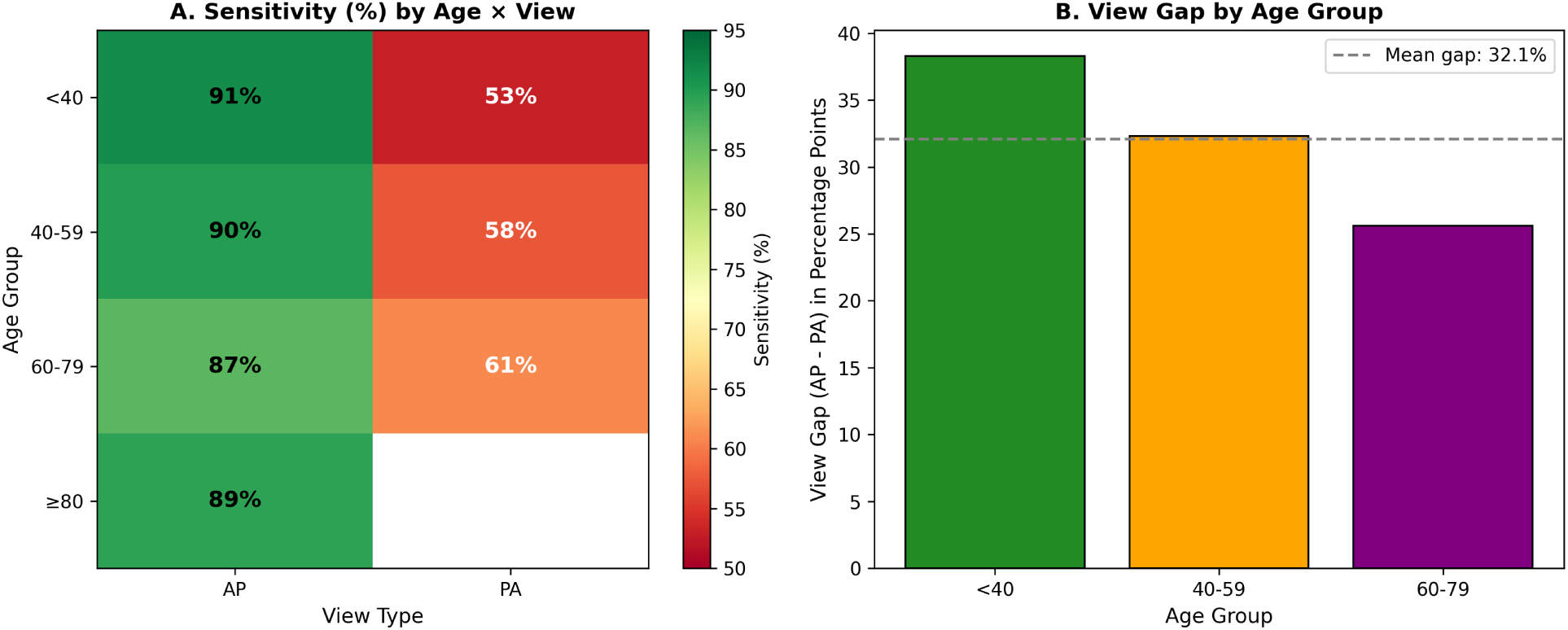
Age × view interaction.

**S12 Fig.**
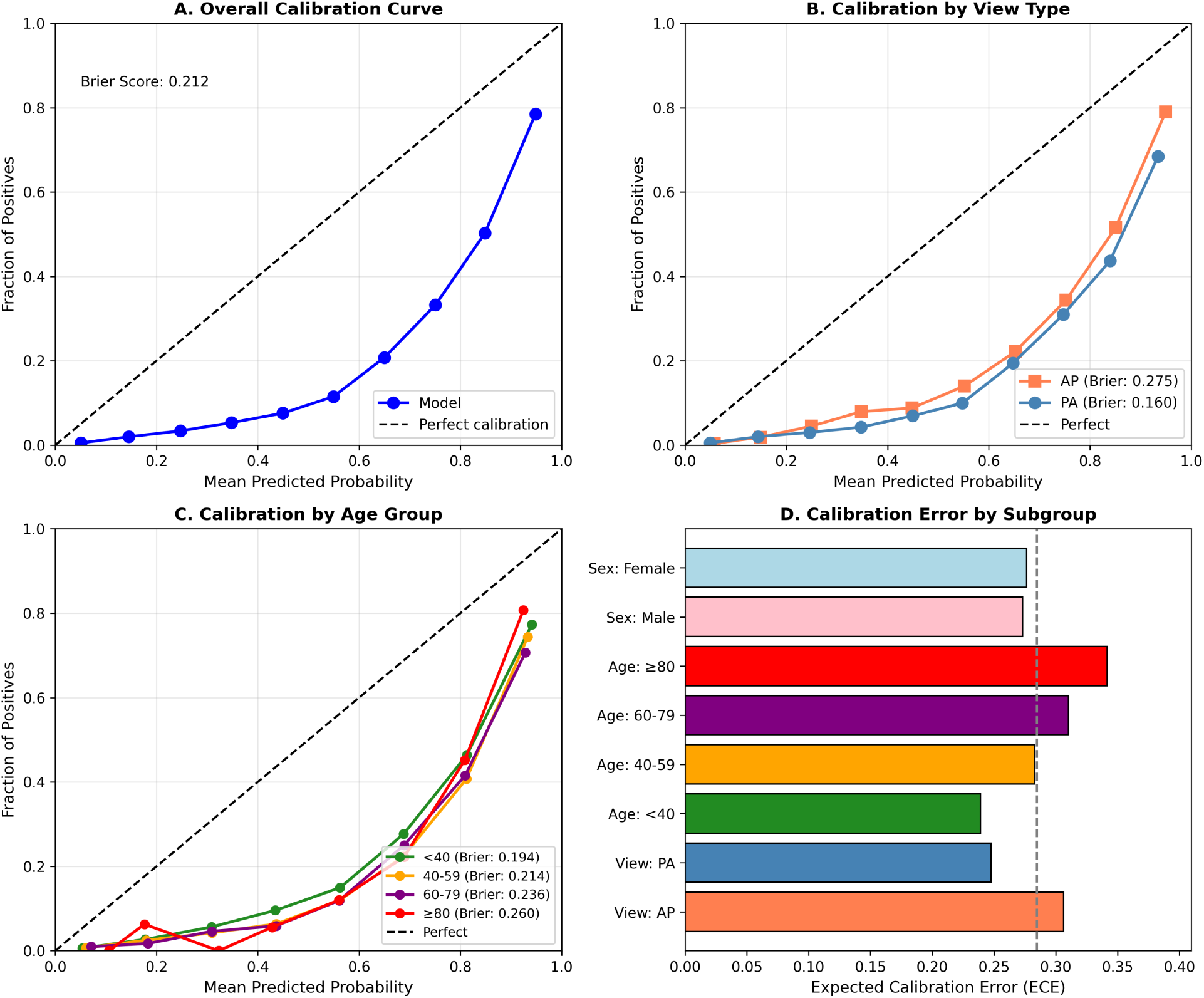
Calibration analysis.

**S13 Fig.**
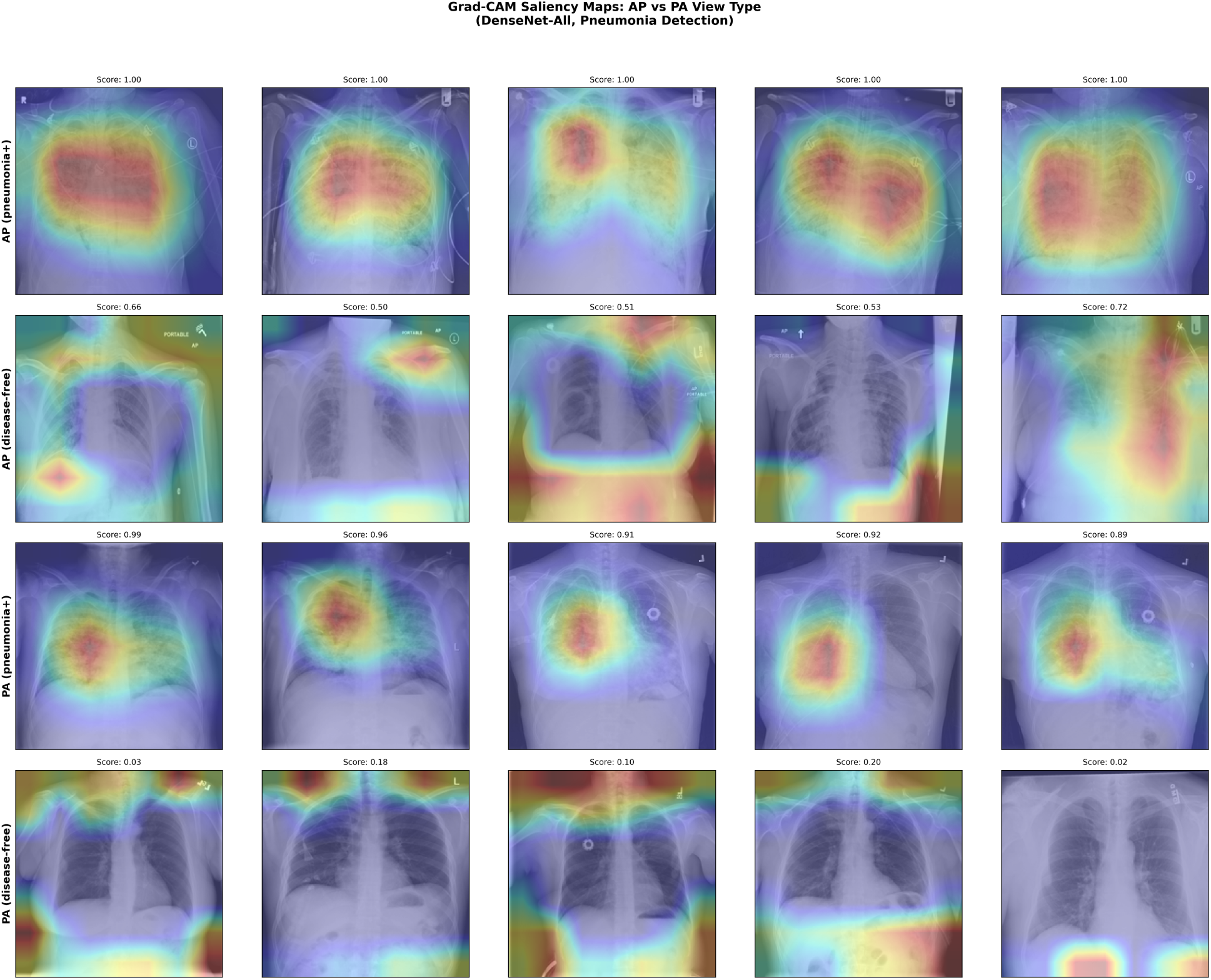
Grad-CAM saliency maps comparing AP vs PA view attention patterns.

